# Novel clinical subphenotypes in COVID-19: derivation, validation, prediction, temporal patterns, and interaction with social determinants of health

**DOI:** 10.1101/2021.02.28.21252645

**Authors:** Chang Su, Yongkang Zhang, James H Flory, Mark G. Weiner, Rainu Kaushal, Edward J. Schenck, Fei Wang

## Abstract

The coronavirus disease 2019 (COVID-19) is heterogeneous and our understanding of the biological mechanisms of host response to the novel viral infection remains limited. Identification of meaningful clinical subphenotypes may benefit pathophysiological study, clinical practice, and clinical trials. Here, our aim was to derive and validate COVID-19 subphenotypes using machine learning and routinely collected clinical data, assess temporal patterns of these subphenotypes during the pandemic course, and examine their interaction with social determinants of health (SDoH). We retrospectively analyzed 14418 COVID-19 patients in five major medical centers in New York City (NYC), between March 1 and June 12, 2020. Using clustering analysis, four biologically distinct subphenotypes were derived in the development cohort (N = 8199). Importantly, the identified subphenotypes were highly predictive of clinical outcomes (especially 60-day mortality). Sensitivity analyses in the development cohort, and re-derivation and prediction in the internal (N = 3519) and external (N = 3519) validation cohorts confirmed the reproducibility and usability of the subphenotypes. Further analyses showed varying subphenotype prevalence across the peak of the outbreak in NYC. We also found that SDoH specifically influenced mortality outcome in Subphenotype IV, which is associated with older age, worse clinical manifestation, and high comorbidity burden. Our findings may lead to a better understanding of how COVID-19 causes disease in different populations and potentially benefit clinical trial development. The temporal patterns and SDoH implications of the subphenotypes may add new insights to health policy to reduce social disparity in the pandemic.

## Introduction

The outbreak of coronavirus disease 2019 (COVID-19), caused by the severe acute respiratory syndrome coronavirus 2 (SARS-CoV-2) infection, has led to a pandemic that imposed tremendous pressure on healthcare systems globally^1^. As the pandemic continues and the second wave has emerged in the US and many other countries, research is still needed to understand how SARS-CoV-2 causes the wide spectrum of COVID-19 disease. Previous studies have uncovered substantial variation in the host response to SARS-CoV-2 and the variable clinical manifestations of this disease, including respiratory failure, kidney injury, and cardiovascular dysfunction^2-8^. Pivotal studies of corticosteroids^9^ and anticoagulation^10,11^ demonstrate differential responses in distinct subpopulations based on severity of disease. The pathophysiology of differential organ dysfunction in COVID-19 remains unclear across varied patient populations. Prior to the COVID-19 pandemic, identification of biologically distinct, data driven subphenotypes^12,13^ has helped to disentangle complex syndromic disease such as sepsis^14,15^, ARDS^16^, heart failure^17,18^, diabetes^19^, and Alzheimer’s disease^20^.

Identifying robust subphenotypes in COVID-19 patients could lead to improved understanding of biological mechanisms of host response to SARS-CoV-2 infection and may identify subpopulations that could be prioritized for clinical trial enrollment^13,21^. Previous efforts^22-25^ have been made in this area but remain limited probably due to cohort size, data availability, and lacking evaluation of robustness and usability of the identified subphenotypes. In addition, the hospitalized case fatality rate of COVID-19 has varied over the course of the pandemic^26,27^ and according to social determinants of health (SDoH)^28-30^. Exploration of temporal patterns and SDoH characteristics in conjunction with subphenotypes may derive new insights to improve public health.

In this analysis, our goal was to derive and validate COVID-19 subphenotypes amongst a population of patients who presented to the emergency department (ED) or were hospitalized in multiple health systems in New York City (NYC). Specifically, we used routinely collected clinical data to first derive subphenotypes using the agglomerative hierarchical clustering model. Then, multiple strategies in data pre-processing, data filtering, and data-driven models (both unsupervised clustering model and supervised predictive model) were used to confirm reproducibility and usability of the identified subphenotypes. After that, statistical analyses were conducted to evaluate the characteristics and clinical outcomes of the subphenotypes. Further analyses were performed to examine temporal patterns of the subphenotypes and impacts of SDoH status on subphenotype-level outcomes. The overall workflow of our study is illustrated in Figure 1.

**Figure 1.**
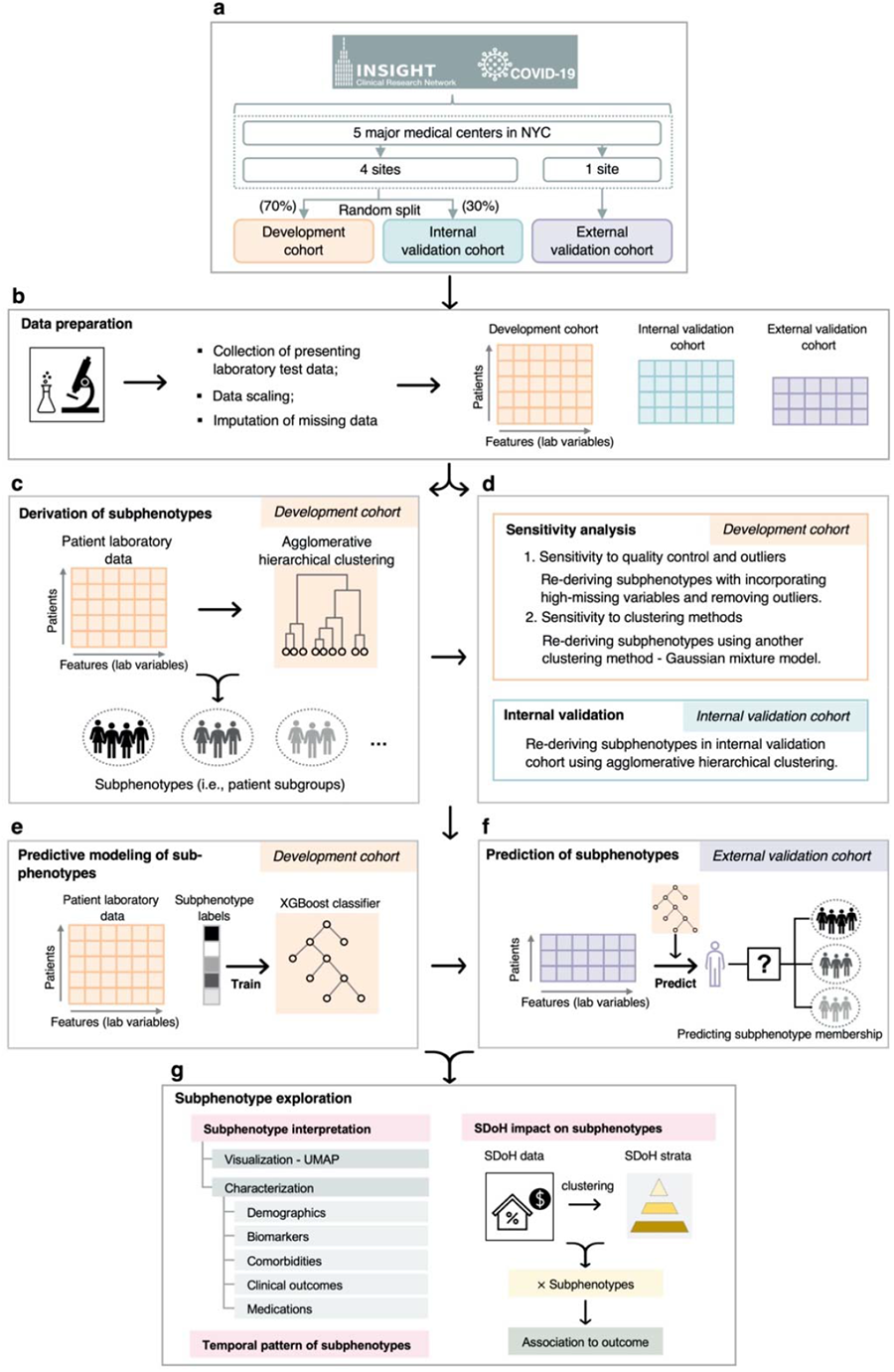
A schematic of the analysis plan. **a**. Strategy for construction of development, internal validation, and external validation cohorts. Studied patients were treated in 5 major medical centers in New York City, including New York University Langone Medical Center, New York Presbyterian - Weill Cornell Medical Center, New York Presbyterian - Columbia University Medical Center, Mount Sinai Health System, and Montefiore Medical Center. **b**. Data preparation for clustering analysis. **c**. Derivation of subphenotypes in the development cohort. Reproducibility of the identified subphenotypes were evaluated in multiple ways, including (**d**) sensitivity analyses in the development cohort and subphenotype re-derivation in the internal validation cohort; and (**e**) training subphenotype predictive model in the development cohort and (**f**) using it to predict subphenotype memberships of patients in the external validation cohort. Last, (**g**) further analyses were conducted to interpret subphenotypes, explore temporal patterns of subphenotypes during the pandemic, and evaluate impact of SDoH characterisitics on subphenotypes. Abbreviations: NYC = New York City; SDoH = social determinants of health; UMAP = Uniform Manifold Approximation and Projection

## Results

### Patients

A total of 14418 patients with confirmed COVID-19 between March 1st and June 12^th^ 2020, treated in ED (N=2354, 16.3%) or inpatient (N=12064, 83.7%) settings, were included for analysis from the five major medical centers in New York City (NYC), including New York University Langone Medical Center (NYU-LMC), New York Presbyterian - Weill Cornell Medical Center (NYP-WCMC), Mount Sinai Health System (MSHS), Montefiore Medical Center (MMC), and New York Presbyterian - Columbia University Medical Center (NYP-CUMC). Details of inclusion and exclusion criteria are presented in eFigure 1 in Supplement. We identified 2853 (19.8%) deaths within 60-day after COVID-19 confirmation in total, including 2801 (19.4%) in-hospital deaths and 52 (4%) deaths after discharge from COVID related hospitalization or ED visits. Considering population diversity (especially race) of the five medical centers (see eTable 1 in Supplement), we combined four centers and randomly divided them into the development cohort (70%) and internal validation cohort (30%); patients of the remaining center were used as the external validation cohort (see Figure 1 and eFigure. 1 in Supplement).

The development cohort contained a total of 8199 patients with a median age of 65.35 (interquartile range [IQR] [50.57, 75.17]) years old, consisting of 3787 (46.2%) females, 2036 (24.8%) white patients, and 2155 (26.3%) black patients. The internal validation cohort contained a total of 3519 patients with similar patient characteristics when compared with the development cohort, with a median age of 63.51 (IQR [50.95, 75,17]) years old, consisting of 1585 (45.0%) females, 838 (23.8%) white patients, and 915 (26%) black patients. The external validation cohort contained a total of 2700 patients. It had a median age of 65.85 (IQR [51.08, 77.38]) years old and consisted of 1305 (48.3%) females, 675 (25.0%) white patients, and 545 (20.2%) black patients. Across the three cohorts, the overall 60-day mortality rates after ED or hospital discharge were 18.65%, 19.78%, and 20.59%, respectively. More details of the characteristics of the studied cohorts appeared in Table 1.

**Table 1.** Characteristics of the development, internal validation, and external validation cohorts.

| Characteristics | Cohort |  |  |
| --- | --- | --- | --- |
|  | Development cohort | Internal validation cohort | External validation cohort |
| No. of patients | 8,199 | 3,519 | 2,700 |
| Construction method | 70% patients (randomly selected) from 4 medical centers | Remaining 30% patients from 4 medical centers | Patients from the last center |
| Age, y, Median (IQR) | 63.53 [50.57 - 75.15] | 63.51 [50.95 - 75.17] | 65.58 (51.08 - 77.39) |
| Sex female, N (%) | 3,787 (46.2) | 1,585 (45.0) | 1,305 (48.3) |
| Race, N (%) |  |  |  |
| White | 2,036 (24.8) | 838 (23.8) | 675 (25.0) |
| Black | 2,155 (26.3) | 915 (26.0) | 545 (20.2) |
| Asian | 409 (5.0) | 193 (5.5) | 28 (1.0) |
| Other/unknown | 3599 (43.9) | 1573 (44.7) | 1452 (53.8) |
| Outcomes (60 days), N (%) |  |  |  |
| Mortality | 1529 (18.65) | 696 (19.78) | 556 (20.59) |
| Mechanical ventilation (intubation) | 1154 (14.07) | 497 (14.12) | 248 (9.19) |
| ICU admission | 1494 (18.22) | 661 (18.78) | - |

Abbreviations: ICU = intensive care unit; IQR = Interquartile range; SDoH = social determinants of health.

### Subphenotypes derivation

In the development cohort, the agglomerative hierarchical clustering model identified 4 distinct subphenotypes based on presenting clinical data of the patients (see eResults, eFigures 3 and 4 in Supplement). Characteristics including demographics, clinical variables, comorbidities, clinical outcomes, and medication treatments across the 4 subphenotypes were presented in Table 2 and Figures 2 and 3.

**Table 2.**
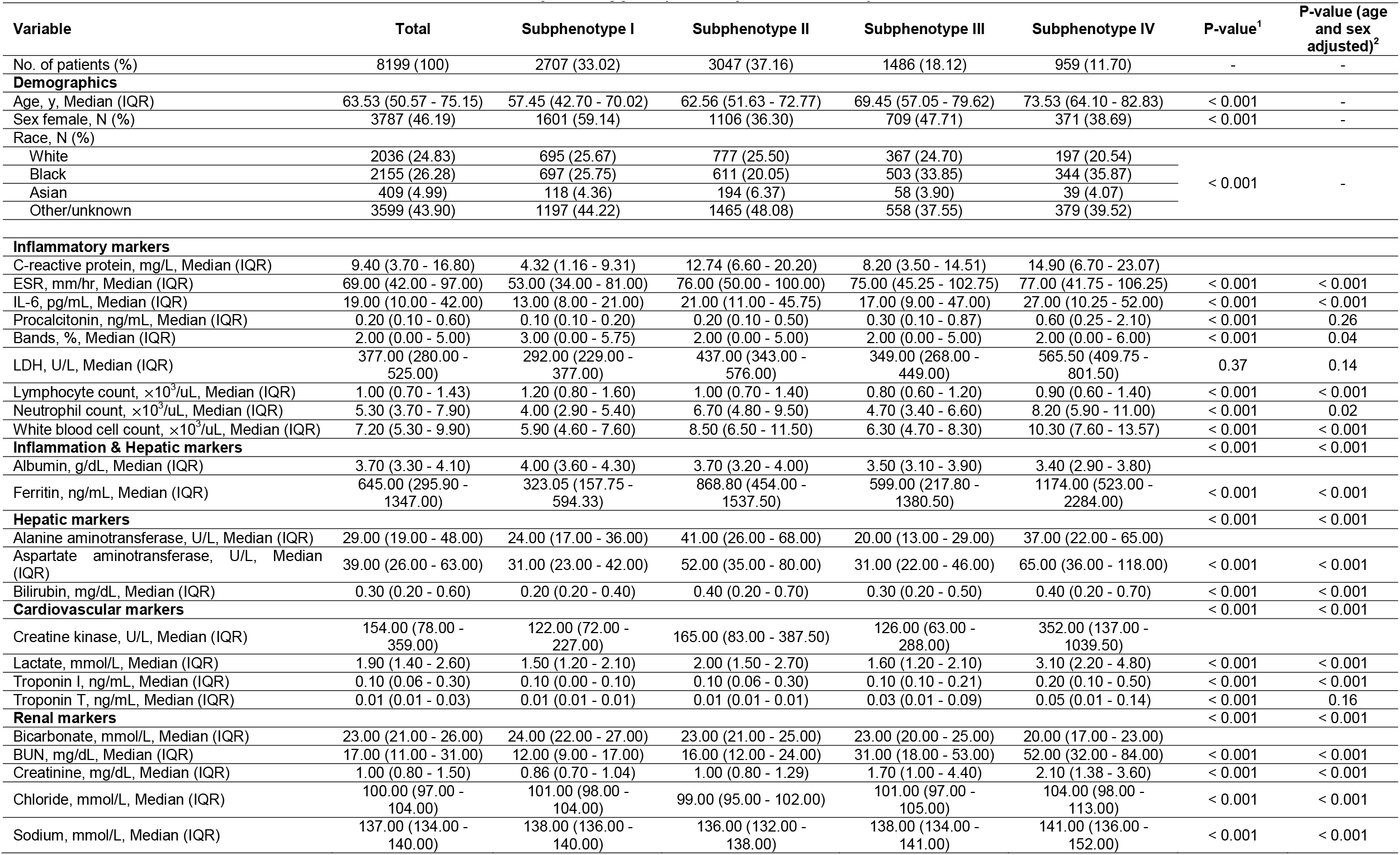
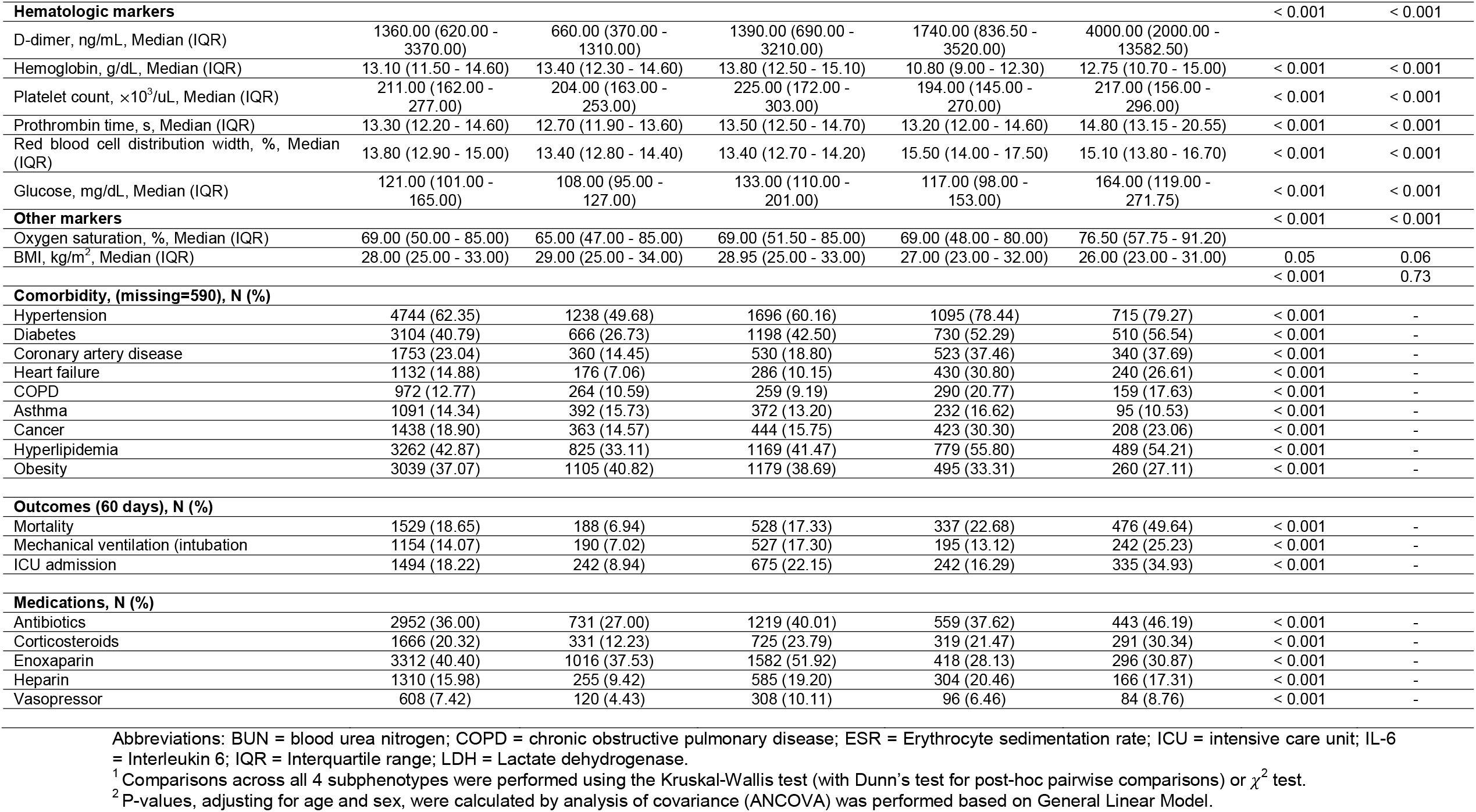
Characteristics of the identified subphenotypes (development cohort)

**Figure 2.**
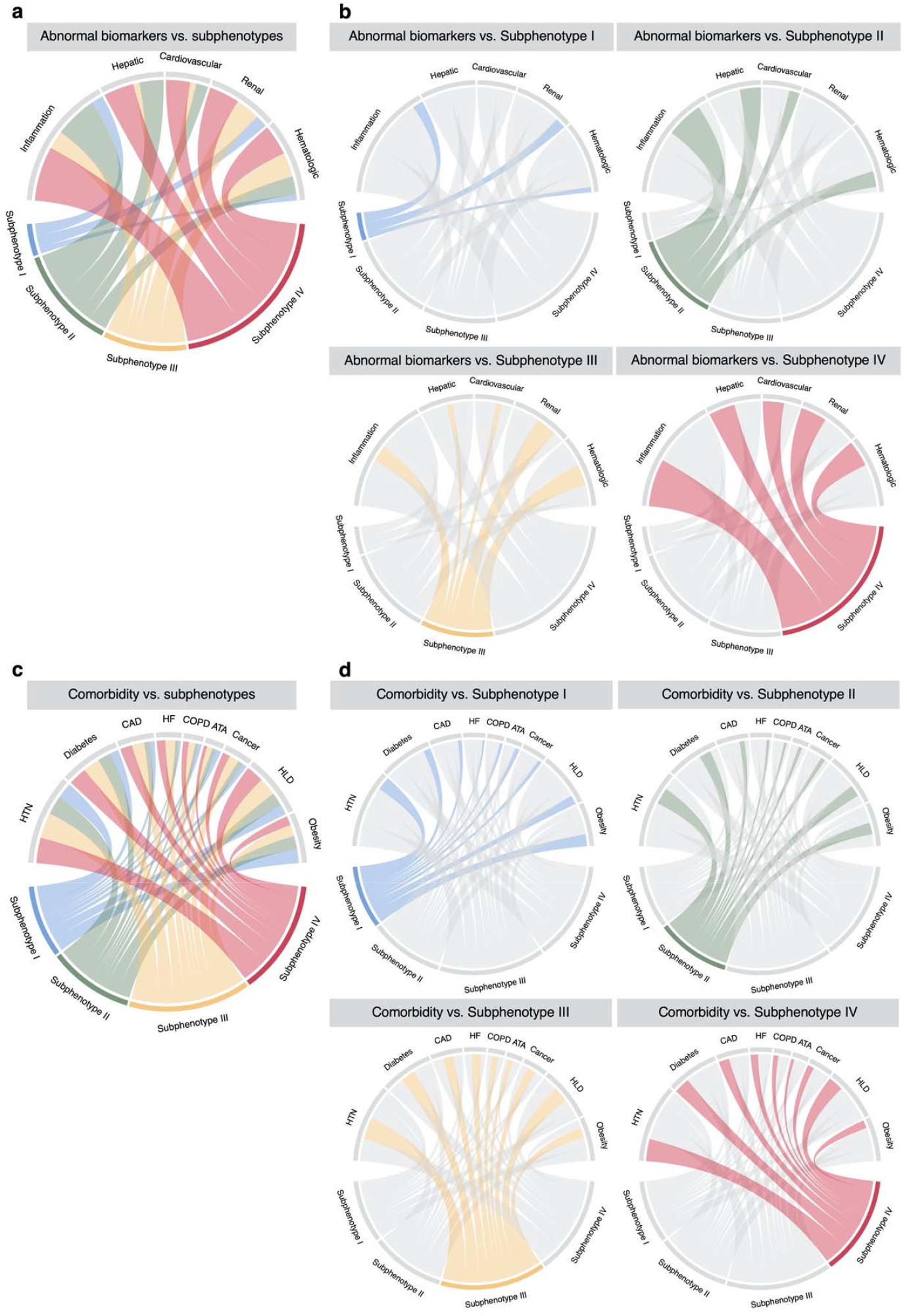
Chord diagrams showing differences in abnormal clinical variables and comorbidity burden among subphenotypes. **a**. Abnormal biomarkers vs. all subphenotypes. **b**. Abnormal biomarkers vs. each subphenotype. **c**. Comorbidities vs. all subphenotypes. **d**. Comorbidities vs. each subphenotype Abbreviations: ATA = asthma; CAD = coronary artery disease; COPD = chronic obstructive pulmonary disease; HF = heart failure; HLD = hyperlipidemia; HTN = hypertension.

**Figure 3.**
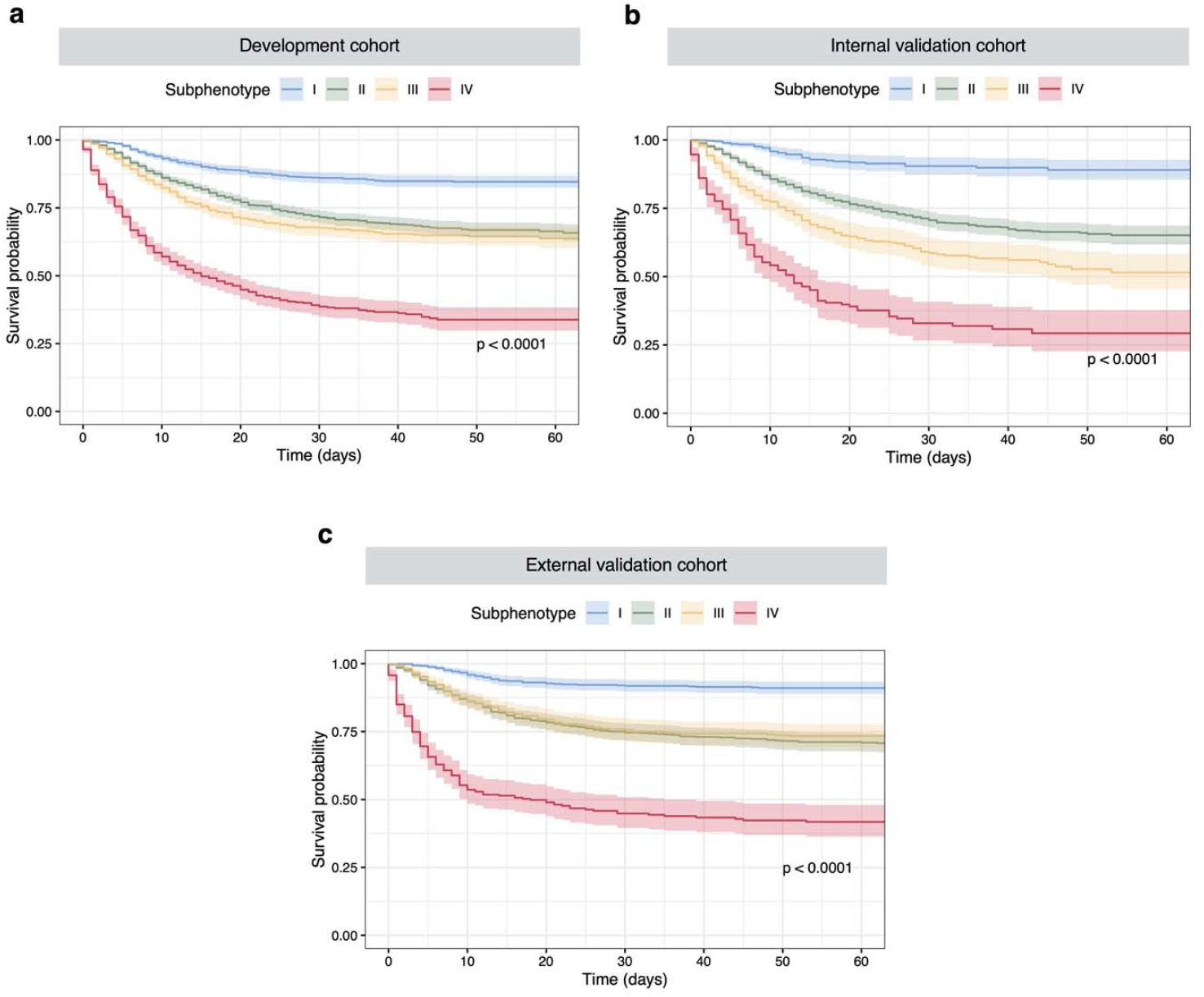
Kaplan-Meier (KM) plots for 60-day mortality by subphenotypes. The survival probabilities were shown with 95% confidence interval. X-axis denotes time (days) after COVID-19 confirmation and Y-axis denotes the survival probability. **a-c**. KM plots by subphenotypes in the development, internal validation, and external validation cohorts, respectively.

#### Subphenotype I

consisted of 2707 (33.02%) patients. Compared to the others, it included more younger (median age 57.45 years, IQR [42.70, 70.02]) and female (N = 1601 [59.15%]) patients. Those patients had more normal values across all clinical variables and lower chronic comorbidity burden. The patients also had better clinical outcomes including a low 60-day mortality (N = 188 [6.94%]) and a low rates of mechanical ventilation (N = 190 [7.02%]) and ICU admission (N = 242 [8.94%]).

#### Subphenotype II

consisted of 3047 (37.16%) patients. Compared to other subphenotypes, it included more male patients (N = 1941 [63.70%]) and was likely to have more abnormal inflammatory markers (such as C-reactive protein, erythrocyte sedimentation rate, interleukin 6, lactate dehydrogenase, lymphocyte count, neutrophil count, white blood cell count, and ferritin) and markers of hepatic dysfunctions (such as ferritin, alanine aminotransferase, aspartate aminotransferase, and bilirubin). Overall comorbidity burden of Subphenotype II was low. Clinical outcomes including 60-day mortality (N = 528 [17.33%]), mechanical ventilation (N = 527 [17.30%]), and ICU admission (N = 675 [22.15%]) of Subphenotype II were at a moderate level.

#### Subphenotype III

included 1486 (18.12%) patients, consisting of more older (median age 69.45 years, IQR [57.05, 79.62]) and black (N = 503 [33.85%]) patients, compared to subphenotypes I and II. Those patients of Subphenotype III were likely to have more abnormal renal dysfunction markers (such as blood urea nitrogen, creatinine, chloride, and sodium) and hematologic dysfunction markers (such as d-dimer, hemoglobin, and red blood cell distribution width). Overall comorbidity burden of Subphenotype III was high. Clinical outcomes including 60-day mortality (N = 337 [22.68%]), intubation (N = 195 [13.12%]), and ICU admission (N =242 [16.29%]) of Subphenotype II were close to that of Subphenotype II and at a moderate level as well.

#### Subphenotype IV

included 959 (11.70%) patients. Compared to other subphenotypes, it included more older (median age 75.53 years, IQR [64.10, 84.83]) and male (N = 588 [61.31%]) patients. Those patients of Subphenotype IV had more abnormal values across all clinical variables and higher chronic comorbidity burden than the others. Obesity burden is lower in Subphenotype IV than others. In line with its biological characteristics, Subphenotype IV had the worst clinical outcomes in 60-day mortality (N = 476 [49.64%]), intubation (N = 242 [25.23%]), and ICU admission (N =335 [34.93%]). In addition, the medications including antibiotics, corticosteroids, and vasopressor were more frequently used in Subphenotype IV.

### Subphenotype reproducibility and prediction

In the development cohort, sensitivity analyses under two different settings (sensitivity to quality control and outliers and sensitivity to clustering methods) confirmed the underlying 4-cluster structure of the data (see eResults, eFigures 3 and 4, and eTable 5 in Supplement). Patients’ memberships of the 4 clusters re-derived by sensitivity analyses were highly consistent with those derived in the primary analysis (see eFigure 6 in Supplement). Moreover, we did not find substantial changes in clinical characteristics of the subphenotypes in the sensitivity analyses (see eTables 6 and 7 in Supplement).

Subphenotypes were also re-derived in the internal validation cohort, where the 4-cluster structure was found as the optimal fit as well (see eResults and eFigure7 in Supplement). Clinical characteristics of the re-derived subphenotypes in the internal validation cohort, including demographics, laboratory variables, comorbidities, and clinical outcomes, also showed very similar patterns with the subphenotypes derived in the primary analysis (see Figure 3, and eTable 8 and eFigure 8 in Supplement).

To further evaluate subphenotype robustness and usability, we trained a predictive model of subphenotypes in the development cohort and used it to predict subphenotype membership in the external validation cohort. Clinical variables of presenting laboratory tests for clustering analysis were used as candidate predictors. The trained predictive model (XGBoost classifier) achieved very high performance in predicting each subphenotype (see eFigure 9 in Supplement). SHapley Additive exPlanation (SHAP) values illustrated contributions of the clinical variables in distinguishing each subphenotype from others (see eFigure 10 in Supplement). Patterns of the produced SHAP values were highly in line with the subphenotype characteristics: 1) normal values of the clinical variables indicated Subphenotype I; 2) abnormal inflammatory and hepatic markers were predictive of Subphenotype II; 3) abnormal renal and hematologic markers indicated were likely to indicate Subphenotype III; 4) Subphenotype IV was associated with abnormal values of most variables. After that, the trained predictive model was used to predict subphenotype memberships of patients in the external validation cohort. The predicted subphenotypes in the external validation cohort were well separated in the UMAP space (see eFigure 11 in Supplement) and showed clinical characteristics similar to findings in the primary analysis (see Figure 3, and eTable 9 and eFigure 12 in Supplement).

Last, results from leave-one-center-out analysis also confirmed the four-cluster structure underlying our data (see eFigure 13 in Supplement). Meanwhile, subphenotypes identified by the leave-one-center-out analysis among the whole population showed characteristics in line with those identified in the primary analysis (see eTable 10 in Supplement). Those above demonstrated stability of the identified subphenotypes across the five centers.

### Temporal characteristics of subphenotypes

Temporal patterns of the COVID-19 subphenotypes were illustrated by the bar charts, showing the composition of subphenotype memberships of patients confirmed per week, since the outbreak in NYC, i.e., March 1, 2020 (see Figures 4a-c). Except week 1 and week 14 that had few patients confirmed, the composition of the four subphenotypes per week evolved over time and showed similar patterns across the development, internal validation, and external validation cohorts. In general, patients with confirmed SARS-CoV-2 infection rapidly increased within the first month since the outbreak and reached the peak at week 5 (early April). Subphenotype I (mild symptom) and Subphenotype II (moderate symptom, low comorbidity burden) dominated the time period prior to the peak (first 4 weeks since outbreak). In contrast, Subphenotype IV (severe symptom, high comorbidity burden) had a low proportion within the first 4 weeks but showed a largely increased proportion from week 6 to week 9. Since week 10, the proportion of Subphenotype I gradually increased while others especially Subphenotype IV shrank. Subphenotype III (moderate symptom, high comorbidity burden) had a relatively stable proportion over time.

**Figure 4.**
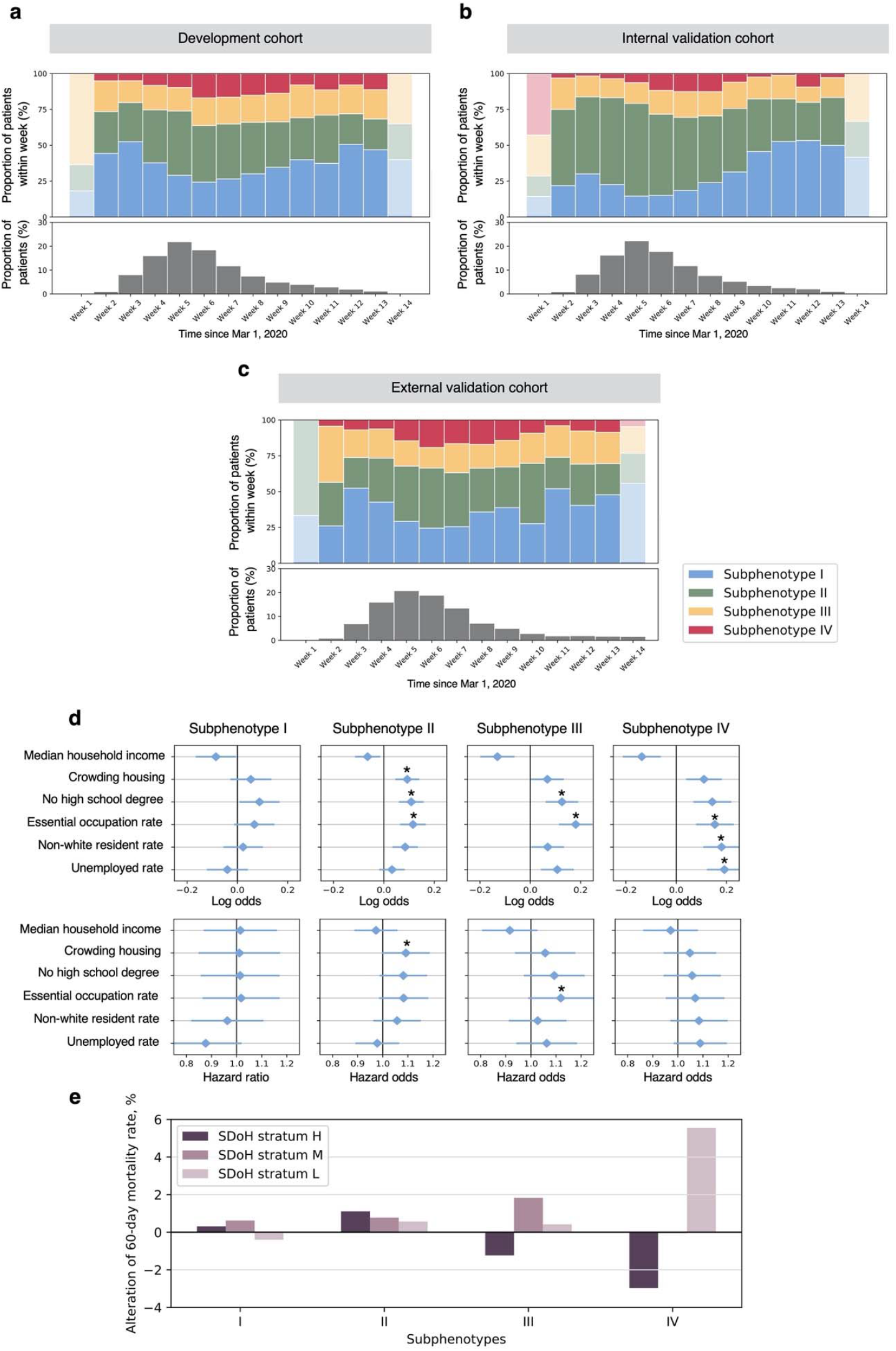
Plots showing temporal patterns and SDoH implications of subphenotypes. **a-c**. Proportions of subphenotype memberships of patients confirmed per week, since March 1, 2020. **d**. Log odds and Hazard ratio (mean values and standard deviation [error bar]) showing associations between individual SDoH characteristics and 60-day mortality risk, using logistic regression analysis and Cox regression analysis, adjusting for age and sex, respectively. **e**. Plot showing alteration of 60-day mortality rate (Y-axis) of each SDoH stratum to that of subphenotype level. * P-value < 0.05

### Impact of SDoH on subphenotypes

In general, worse SDoH in terms the socioeconomic variables were likely in Subphenotype IV (see eTable 11 in Supplement). Moreover, logistic regression analysis identified similar patterns of relationships between the SDoH variables with 60-day mortality risk across subphenotypes; however, absolute log odds and Hazard ratio of the SDoH variables varied across subphenotypes (see Figure 4d and eTables 12-13 in Supplement). For example, low absolute log odds were observed in all six SDoH variables in Subphenotype I. In contrast, we did see increased absolute log odds of all six SDoH variables in Subphenotype IV. Hazard ratio showed similar pattern.

Agglomerative hierarchical clustering based on the SDoH variables grouped the patients into a 3-cluster model (see eResults and eFigure 14 in Supplement), which can be interpreted as high (H), middle (M), and low (L) SDoH strata (see eTable 14 in Supplement). Stratum L, representing disadvantaged SDoH status, accounted for a slightly higher mortality rate (H vs. M vs. L, 17.59% vs. 19.91% vs.19.98%, P-value = 0.08). In addition, stratum L had a lower ICU admission rate (16.16%, P-value < 0.001). The relative high mortality but low ICU admission rate may be caused by critical care strain during periods of increased COVID-19 ICU demand, as suggested by a recent study^31^. Distributions of the SDoH strata by biological subphenotypes were shown in eTable 15 in Supplement. In the analysis to further explore how SDoH strata affected the outcome of each biological subphenotype, we found varied patterns of correlations between SDoH strata and 60-day mortality (see Figure 4e) by subphenotypes. Notably, in line with the results of the univariate analysis above, SDoH strata were likely to have a strong impact on the 60-day mortality in Subphenotype IV. Particularly, in Subphenotype IV, SDoH stratum L was associated with a 55.19% 60-mortality rate, which was 5.55% higher than the subphenotype level (49.64%, see Table 2) and 8.52% higher than that of the SDoH stratum H. In subphenotypes I, II, and III, we didn’t find mortality rate discrepancy higher than 3% between any pair of SDoH strata. Similarly, considering stratum H as reference, stratum L had largely increased log odds of mortality in Subphenotype IV (log odds = 0.40, SD = 0.19, P-value = 0.04). (see eTable 16 in Supplement)

## Discussions

We derived subphenotypes of COVID-19 patients treated at five major medical centers in NYC across the whole course of the first wave of the pandemic, using the clinical data at the presentation to the emergency department (ED) or hospital. Different from the previous subphenotype studies of COVID-19^22-24^, we focused on a larger, more representative, and diverse population presented at the ED and/or hospitalized without COVID-19 specific therapy. We derived subphenotypes using clustering analysis in the development cohort and validated them using a combination of multiple validation strategies, including the use of different data processing, different data filtering, and different machine learning models (both unsupervised clustering and supervised predictive models). All validation approaches confirmed the reproducibility of the 4-cluster structure of the data and clinical characteristics of the identified subphenotypes. We would also highlight that all machine learning models used for subphenotype derivation and validation were performed only on the presenting clinical variables that were routinely collected in daily patient care and are available to providers by ED or hospital admission. This allows us to potentially capture the underlying variable mechanisms of the complex disease, but also enhances the generalizability and feasibility of the identified subphenotypes to be used in clinical practices and patient enrollment in clinical trials.

Importantly, the 4 subphenotypes identified were significantly separated in demographics, clinical variables, and chronic comorbidities, and strongly predictive of the 60-day mortality outcome. Subphenotype IV included more older, male patients, abnormal markers indicating hyperinflammation, liver injury, cardiovascular problems, renal dysfunctions, and coagulation disorders, and a higher comorbidity burden (except for obesity) compared to the other subphenotypes. In contrast, Subphenotype I was composed of relatively healthy, younger females who had more normal values across all markers and comorbidity burdens compared to the other subphenotypes. There was a strong concordance between their clinical profiles and outcomes, such as Subphenotype IV showed the worst clinical outcome while Subphenotype I showed the best outcome among the 4 subphenotypes. These are in line with observations reported in a previous small cohort study^23^. Interestingly, Subphenotypes II and III showed similar, moderate-level 60-day mortality rates, but their clinical characteristic profiles suggested that they were likely to have distinct biological mechanisms. In particular, results from our primary analysis and validation approaches demonstrated that Subphenotype II was correlated with relative hyperinflammation, while Subphenotype III was associated with renal injury, lower platelet level and a high comorbidity burden (significantly higher than Subphenotypes I and II, and equivalent to Subphenotype IV). Moreover, in accordance with the clinical characteristics and outcomes, the worse subphenotypes (Subphenotypes III and IV) were more likely to receive medications in antibiotics, corticosteroids, and vasopressor than the others. These findings suggested that our identified subphenotypes offer insight into the varied mechanisms of COVID-19.

Typically, data-driven approaches for the identification of subphenotypes of human disease are based on the unsupervised clustering methods^12,14-16,22-24,32^. The natural attributes of the unsupervised methodology in discovering underlying patterns from data make them the best fit for subphenotype identification. Once the subphenotypes were determined, there would be a need of subphenotype membership assignments for new patients. However, previous studies barely discussed such down-stream usability of the identified subphenotypes. In this analysis, we built a supervised predictive model of the identified subphenotypes. Our predictive model achieved an ideal prediction performance in the development cohort and predicted subphenotypes in the external validation cohort that presented the same pattern of clinical characteristics with that of the originally derived subphenotypes. In this way, instead of validating the subphenotypes in a different route, the predictive model brought additional implications as: 1) it provides a feasible and accurate way to apply the identified subphenotypes to clinical practice; and 2) contributions of the clinical variables in subphenotype prediction calculated by the SHAP method showed concordant patterns with the subphenotypes’ clinical characteristics and hence confirmed biological profiles of the subphenotypes in the multivariate prospective.

Time is a crucial factor in the spread of COVID-19. Previous studies have examined the temporal trends of COVID-19 outcomes such as in-hospital mortality rate during the course of the pandemic^26,27^, but limited attention has been drawn on evolving patterns of COVID-19 phenotypes. We filled this gap in the present study. Our observations suggested varied temporal trends of the identified subphenotypes during the first 14 weeks of the pandemic in NYC. Interestingly, since the COVID-19 outbreak in NYC on March 1, 2020, Subphenotypes I and II dominated the time period prior to the peak (first 4 weeks since outbreak), possibly as they contained more relatively younger patients who may have had more frequent social activities to be infected. Subphenotype IV, with older age, worse health conditions, and poorer outcomes, was boosted within the second month (April 2020) post spread peak, consistent with tremendous mortality rate of NYC in April^33^.

This would suggest that younger, biologically strong patients (Subphenotypes I and II) got infections early and boosted the spread, while older, biologically vulnerable patients (Subphenotype IV) accounted for the second infections within a population probably due to housing. After that, the proportion of Subphenotype I out of all patients confirmed per week gradually expanded while that of the others, especially Subphenotype IV shrank. The potential reason would be that valuable experience (such as the improved use of masks and social distancing), reinforced health care systems, and announced health policies did protect the population who likely develop severe subphenotypes (Subphenotype IV). In general, such temporal trends of the biological subphenotypes would be a considerable, fine-grained explanation of the observed outcome (mortality rate) evolving trends in epidemiology^26^.

SDoH such as vulnerable socioeconomic neighborhood status have been associated with poor outcomes of COVID-19^26,30^. In this work, we explored the impact of SDoH on different biological subphenotypes from both univariate and multivariate perspectives. We first examined the associations of individual socioeconomic characteristics with mortality risk in each subphenotype. We then derived comprehensive SDoH strata using the data-driven clustering method and evaluated their correlations with mortality risk in each subphenotype. The results confirmed our hypothesis that SDoH impacts biological subphenotypes differently. The highly expanded mortality risk log odds of individual SDoH variables and discrepancy of mortality rate among SDoH strata indicate that SDoH has a much stronger association with mortality outcomes in Subphenotype IV, compared to the others. In other words, once a sick, elderly patient shows up with COVID-19 (Subphenotype IV), the disadvantaged socioeconomic status significantly increased their mortality. In contrast, disadvantaged SDoH status was unlikely to lead to significantly increased mortality risk in Subphenotype I. This evidence further demonstrated that the COVID-19 pandemic has disproportionately affected patients with lower socioeconomic status. In general, our findings added new information on social disparities in the COVID-19 pandemic. Unlike previous studies^29,30,34,35^ that focused on the entire population, we extended the study from a new angle by focusing on the biologically different populations (i.e., subphenotypes). Our findings also showed evidence that the identified subphenotypes would provide considerable guidance in health policy to reduce social disparities in the pandemic.

### Limitations

While this study presents a new contribution in the efforts to parse the biological heterogeneity of COVID-19, there remain several limitations. First of all, our data-driven approach relied on the availability of patient data. In this study, we identified subphenotypes using the routinely collected clinical variables that were correlated with COVID-19^36^ and available in the INSIGHT database^37^. We were not able to extract presenting symptoms and vital data while the incorporation of such data would add in new insights.

Second, in our study, the analyzed data were collected at ED or hospital presentation, so the time between COVID-19 symptom onset to ED or hospital presentation could be a covariate of disease severity and clinical outcomes. However, such data was not available in the INSIGHT database.

Third, missing values may affect the robustness of the identified subphenotypes. In order to address this issue, we excluded variables with high missingness. For the remaining variables, we used the K-nearest neighbors imputation algorithm^38^. Even so, we still missed these real values hence may incorporate bias.

Fourth, our study was based on presenting clinical data, such that each patient was characterized in a snapshot. The full use of longitudinal data of patients may allow us to capture the complexity of the disease arc to identify interesting subphenotypes. Previous studies tried to derive COVID-19 subphenotypes based on longitudinal information^22,24^, yet they were based on univariate trajectory data in small cohorts. The collection of multivariate, longitudinal data in large cohorts remains challenging and modeling such data to identify subphenotypes requires improved data-driven methods^12,13,21^.

Fifth, this is a multiple institutional analysis in NYC. To evaluate the generalizability of the identified subphenotypes, further validation on data collected from other areas is needed in future work.

## Methods

### Study design and cohort description

We used data of COVID-19 patients from INSIGHT Clinical Research Network (CRN)^37^. INSIGHT is funded by the Patient-Centered Outcomes Research Institute (PCORI) and aggregates clinical data of diverse patient populations across five academic medical centers in New York City (NYC), including New York University Langone Medical Center (NYU-LMC), New York Presbyterian - Weill Cornell Medical Center (NYP-WCMC), New York Presbyterian - Columbia University Medical Center (NYP-CUMC), Mount Sinai Health System (MSHS), and Montefiore Medical Center (MMC). COVID-19 diagnosis was defined as having at least one positive laboratory test result for SARS-CoV-2 infection or at least one ICD-10 diagnosis code for COVID-19 (see eMethods in Supplement). Study participants were adult patients who were diagnosed with COVID-19 and treated in ED or inpatient settings in these five health centers from March 1 to June 12, 2020. Criteria used to assess patient eligibility are illustrated in eFigure 1 in Supplement. Exclusion criteria include younger than 18 years old; duplicated patient IDs; having no emergency department (ED) or inpatient (IP) admission within 14 days after COVID-19 confirmation; or having missing values on all clinical variables. Considering the population diversity of the five medical centers (see eTable 1 in Supplement), we combined patients of four centers and randomly divided them into the development cohort (70%) and internal validation cohort (30%). Patients of the last center were used as the external validation cohort.

### Candidate variables for subphenotype identification

We considered 30 clinical variables associated with COVID-19 onset, symptoms, or outcomes^36^ and available in the INSIGHT database as the candidate variables to derive subphenotypes. The variables included inflammatory markers (C-reactive protein, erythrocyte sedimentation rate [ESR], interleukin 6 [IL-6], procalcitonin, bands [i.e., premature neutrophil], lactate dehydrogenase [LDH], lymphocyte count, neutrophil count, and white blood cell count), inflammatory and hepatic markers (albumin and ferritin), hepatic markers (alanine aminotransferase [ALT], aspartate aminotransferase [AST], and bilirubin), markers of cardiovascular conditions (creatine kinase [CK], lactate, troponin I, and troponin T), markers of renal dysfunctions (bicarbonate, blood urea nitrogen [BUN], creatinine, chloride, and sodium), markers of hematologic dysfunctions (d-dimer, hemoglobin, platelet count, prothrombin time [PT], red blood cell distribution width [RDW], and glucose), and oxygen saturation. For each patient, we extracted the first value of each clinical variable within the collection window, which was defined as: 1) time period from COVID-19 confirmation to the first inpatient encounter, if the patient has an inpatient admission within 14 days after confirmation; or 2) 14 days after COVID-19 confirmation if there was only ED encounters but no inpatient admissions following the COVID-19 diagnosis. If there was no record in the collection window, we extracted the last value within 3 days before confirmation (see eFigure 2 in Supplement).

### Other clinical characteristics, clinical outcomes, and medications

We also examined other clinical characteristics of the patients, including demographics, comorbidities, and body mass index (BMI). Demographics included age, sex, and race. Baseline comorbidities included hypertension, diabetes, coronary artery disease (CAD), heart failure, chronic obstructive pulmonary disease (COPD), asthma, cancer, obesity, and hyperlipidemia. For each patient, the most recent BMI data was collected. We analyzed 60-day all-cause mortality as the primary outcome for the patients. Need for mechanical ventilation and admission to the intensive care unit (ICU) were the secondary outcomes. We also analyzed the treatments for COVID-19, including antibiotics (combining ceftriaxone, azithromycin, piperacillin tazobactam, meropenem, vancomycin, and doxycycline), corticosteroids (combining prednisone, methylprednisolone, dexamethasone, and hydrocortisone), hydroxychloroquine, enoxaparin, heparin, and vasopressor. These above data were collected from patient records available in the INSIGHT database as well.

### SDoH data

To explore the impact of SDoH status on the subphenotypes, we extracted patients’ neighborhood socioeconomic characteristics, including median household income, percentage of residents without a high school degree, percentage of residents who are essential workers, percentage of households with crowding housing conditions (i.e., households with >1 person per room), percentage of non-white residents, and unemployment rate. These characteristics were extracted from the 2018 American Community Survey^39^. Previous studies^40-46^ have indicated that these social conditions are associated with higher probability of infection, hospitalization, and other adverse outcomes, e.g., mortality, in COVID-19.

### Statistical methods

#### Data preparation

We first assessed the value distributions and missingness of the 30 candidate clinical variables (see eTables 2 and 3 in Supplement). For data quality control, 7 variables of high missingness (missing more than 70% values) were excluded and the remaining 23 variables were used for deriving subphenotypes. Logarithmic transformation was applied to the non-normal distributed variables (see eTable 4 in Supplement). In order to eliminate the effects of value magnitude, all variables were scaled based on z-score. K-nearest neighbors (KNN) imputation^38^ was used to address missing values (see eMethods in Supplement).

#### Subphenotype derivation, validation, and prediction

We originally derived subphenotypes using the development cohort. More specifically, agglomerative hierarchical clustering with Euclidean distance calculation and Ward linkage criterion^47^ was applied to the 23 clinical variables after data preparation. We used agglomerative hierarchical clustering because it is robust to different types of data distributions and typically produces a dendrogram that visualizes data structure to help determine the optimal cluster number. Besides dendrogram, we calculated 21 measures of clustering models provided by ‘NbClust’ software^48^ to determine the optimal number of clusters, i.e., subphenotypes. In order to evaluate the reproducibility, we validated our subphenotypes in four ways. First, we performed sensitivity analyses using the development cohort to evaluate 1) sensitivity to quality control and outliers and 2) sensitivity to clustering algorithms. To assess sensitivity to quality control and outliers, we incorporated all 30 candidate variables and excluded patients who have outlier values (see eMethods in Supplement). Then similar to the primary analysis, we performed agglomerative hierarchical clustering to re-derive subphenotypes and determined optimal cluster number using dendrogram and ‘NbClust’. To assess sensitivity to clustering algorithms, we re-derived subphenotypes using the Gaussian mixture model (GMM)^49^, which is a probabilistic model for clustering analysis based on a mixture of Gaussian distributions. The optimal cluster number in GMM was determined by comprehensively considering Akaike information criterion (AIC), Bayesian information criterion (BIC), and median probability of group membership (see eMethods in Supplement).

Second, we used the internal validation cohort and re-derived subphenotypes using the same agglomerative hierarchical clustering with the primary analysis for validation. The optimal cluster number was determined using dendrogram and ‘NbClust’ as well.

Third, for the aims of confirming subphenotypes and their usability, we used the supervised predictive model. More specifically, considering subphenotype membership of each patient as the label to predict, we built a predictive model of subphenotypes based on the 23 clinical variables used for subphenotype derivation. The predictive model was based on the supervised XGBoost classifier^50^, a powerful tree-based machine learning model. The predictive model was trained in the development cohort using a 10-fold cross-validation strategy. To address the multi-label classification (since we identified more than 2 subphenotypes), a one-vs-the-rest strategy was used in model training. Prediction performance was measured by receiver operating characteristics curve (ROC) and area under ROC curve (AUC). We also engaged the

SHapley Additive exPlanation (SHAP) values to assess contributions of the clinical variables in distinguishing each subphenotype from the others. Once the predictive model was trained, it was performed on the external validation cohort to predict the patients’ subphenotype memberships.

Last, to assess stability of the subphenotypes across the five medical centers, we further performed leave-one-center-out analysis (see eMethods in Supplement).

#### Subphenotype interpretation

For the aim of subphenotype interpretation, we first visualized the subphenotypes in two ways: 1) 2-D visualization calculated by Uniform Manifold Approximation and Projection (UMAP) algorithm^51^ based on clinical variables for clustering (showing distributions of subphenotypes within low-dimensional space); 2) chord diagrams^52^ showing differences of subphenotypes in terms of abnormal clinical variable groups and comorbidities (see eMethods in Supplement).

We also characterized subphenotypes by evaluating their differences in demographics, all clinical variables, comorbidities, clinical outcomes, and medications prescribed after COVID-19 confirmation. Data were presented as median (interquartile range [IQR]) for continuous variables and exact patient number (percentage) for categorical variables. To compare subphenotypes, we performed the Kruskal-Wallis test for continuous data and *χ*^2^ test for categorical data. Analysis of covariance (ANCOVA) was also applied for between-subphenotypes comparisons, adjusting for age and gender. Two-tailed P-values smaller than 0.05 were considered as the threshold for statistical significance. Survival analyses were performed to assess associations of subphenotypes to clinical outcomes, where Kaplan-Meier plots were created accordingly.

#### Temporal pattern of subphenotypes

To evaluate the temporal pattern of the subphenotypes during the course of the pandemic, we created bar charts to visualize the proportion of each subphenotype out of the total patients confirmed per week, since the COVID-19 outbreak in NYC (March 1, 2020).

#### Impacts of SDoH on COVID-19 subphenotypes

Multiple analyses were conducted to assess the impact of SDoH on COVID-19 subphenotypes. For each subphenotype, we first performed logistic regression analysis and Cox regression analysis to assess the association of each SDoH variable with 60-day mortality, adjusting for age, sex, and/or clinical variables. After that, we performed agglomerative hierarchical clustering on the 6 socioeconomic variables to derive comprehensive SDoH strata. Within each subphenotype, we compared 60-day mortality rates between the SDoH strata. We also used logistic regression analysis and Cox regression analysis to assess the association of SDoH strata with 60-day mortality, adjusting for age and sex, within each subphenotype.

## Data Availability

All data studied in this work can be downloaded from INSIGHT clinical research network at https://insightcrn.org/our-data/, via request. Implementation of our work is based on Python 3.7 and R 3.6. More specifically, clustering models were implemented based on Python packages scikit-learn 0.23.2 (https://scikit-learn.org/stable/) and scipy 1.5.3 (https://www.scipy.org). Supervised predictive modeling was based on XGBoost 1.2.1 (https://xgboost.readthedocs.io/en/latest/) and SHAP 0.35.0 (https://shap.readthedocs.io/en/latest/). Data dimension reduction and visualization were performed based on Python package UMAP-learn 0.3.9 (https://umap-learn.readthedocs.io/en/latest/). R package NbClust (https://cran.r-project.org/web/packages/NbClust/NbClust.pdf) was used to calculate measures of clusters to determine the optimal cluster number in agglomerative hierarchical clustering. Chord diagrams were created using R package circlize (https://cran.r-project.org/web/packages/circlize/index.html). All statistical tests and survival analyses were performed based on R.

## Ethical approval and patient consent

The Institutional Review Board of the Weill Cornell Medicine approved this study (Protocol number: 20-04021948).

## Data availability

All data studied in this work can be downloaded from INSIGHT clinical research network at https://insightcrn.org/our-data/, via request.

## Code availability

All computer codes in this study are available at https://github.com/ChangSu10/COVID-Insight-subphenotyping. Implementation of our work is based on Python 3.7 and R 3.6. More specifically, clustering models were implemented based on Python packages ‘scikit-learn 0.23.2’ (https://scikit-learn.org/stable/) and ‘scipy 1.5.3’ (https://www.scipy.org). Supervised predictive modeling was based on ‘XGBoost 1.2.1’ (https://xgboost.readthedocs.io/en/latest/) and ‘SHAP 0.35.0’ (https://shap.readthedocs.io/en/latest/). Data dimension reduction and visualization were performed based on Python package ‘UMAP-learn 0.3.9’ (https://umap-learn.readthedocs.io/en/latest/). R package ‘NbClust’ (https://cran.r-project.org/web/packages/NbClust/NbClust.pdf) was used to calculate measures of clusters to determine the optimal cluster number in agglomerative hierarchical clustering. Chord diagrams were created using R package ‘circlize’ (https://cran.r-project.org/web/packages/circlize/index.html). All statistical tests and survival analyses were performed based on R.

## Acknowledgements

This study is funded by the COVID-19-Related Project Enhancement to the grant PCORI/HSD-1604-35187 (“Identifying and Predicting Patients with Preventable High Utilization”, PI: Kaushal) from the Patient-Centered Outcomes Research Institute.

## Author contributions

E.S. and F.W. for conceptualization, investigation, writing, reviewing and editing of the manuscript. C.S. for investigation, data analysis, drafting, editing and reviewing manuscript. M.G.W. for providing data support, discussion and commenting the manuscript. Y.Z., J.H.F., and R.K. for discussion, commenting and editing the manuscript.

