## Supplemental file for "Novel clinical subphenotypes in COVID-19: derivation, validation, prediction, temporal patterns, and interaction with social determinants of health"

### eMethods

#### A. COVID-19 definition

COVID-19 were defined as either having at least one positive laboratory test result or at least one ICD-10 diagnosis code available in INSIGHT database, including:

- LOINC code 94500-6, SARS coronavirus 2 RNA [Presence] in Respiratory specimen by NAA with probe detection;
- LOINC code 94309-2, SARS coronavirus 2 RNA [Presence] in Unspecified specimen by NAA with probe detection;
- LOINC code 94507-1, SARS coronavirus 2 IgG Ab [Presence] in Serum or Plasma by Rapid immunoassay;
- LOINC code 94508-9, SARS coronavirus 2 IgM Ab [Presence] in Serum or Plasma by Rapid immunoassay;
- LOINC code 94306-8, SARS coronavirus 2 RNA panel - Unspecified specimen by NAA with probe detection;
- ICD-10 code B34.2, Coronavirus infection, unspecified site;
- ICD-10 code B97.21, SARS-associated coronavirus as the cause of diseases classified elsewhere;
- ICD-10 code B97.29, Other coronavirus as the cause of diseases classified elsewhere;
- ICD-10 code J12.81, Pneumonia due to SARS-associated coronavirus;
- ICD-10 code U07.1, COVID-19, virus identified.

#### B. Data pre-processing and missing value imputation

Data distributions and missing values of the studied cohorts were assessed (see eTables 2 and 3 in Supplement). 7 out of 30 candidate clinical variables were excluded for cluster analysis for subphenotype derivation, due to high missing rate (missing more than 70% values). The 7 high-missing variables were included in sensitivity analysis. Details of usage of the clinical variables were presented in Table 4 in Supplement.

Logarithmic transformation was applied to the non-normal distributed variables (see eTable 4 in Supplement). In order to eliminate the effects of value magnitude, all variables were scaled based on z-score. More specifically, in the development cohort, z-score is calculated by

$$x' = \frac{x - \mu}{\sigma}$$

where $x$ is the raw value, and $\mu$ and $\sigma$ are the mean and standard deviation of the samples. For each validation cohort, z-score is calculated by:

$$x' = \frac{x - \mu_{dev}}{\sigma_{dev}}$$

where $\mu_{dev}$ and $\sigma_{dev}$ are the mean and standard deviation of the samples of the development cohort.

To address missing values, we used the K-nearest neighbors (KNN) imputation algorithm, which typically selects top K samples most similar to the sample of interest to impute missing values[1]. According to the previous study[1]. we set K=10 in this analysis. Of note, the KNN imputation model was fitted in development cohorts, and then used to impute missing values in development, internal validation, and external validation cohorts.

**C. Agglomerative hierarchical clustering**

In this study, agglomerative hierarchical clustering analysis was used to derive subphenotypes. On one hand, unlike other clustering methods like k-means clustering, agglomerative hierarchical clustering is usually robust as it’s not sensitive to data distribution (e.g., k-means requires a sphere-like distribution of the data) and doesn’t need an initialization procedure that may incorporate uncertainty. On the other hand, agglomerative hierarchical clustering typically produces a tree diagram known as dendrogram, which visually illustrates how the data points are agglomerated together in a hierarchical manner and distances between the clusters at different layers in the hierarchy, providing visible guidance in determining the optimal cluster number. For example in eFigure 3a, each data point (i.e., patient) is considered as a separate cluster at the beginning. Then, at each step, the two clusters that are most similar are joined into a single new cluster. The vertical axis of the dendrogram represents the distance or dissimilarity between clusters. High inter-cluster distance indicates a clear cluster structure of the data. Here, we performed agglomerative hierarchical clustering with Euclidean distance calculation based on clinical variable of the patients. Ward linkage criterion[2] was used to construct the hierarchy.

'NbClust'[3] is a famous R package that was developed to assist a clustering method to determine the optimal cluster number of the data. In this study, we used 21 indices provided by 'NbClust' to evaluate cluster structure of the agglomerative hierarchical clustering model with Ward criterion. The optimal cluster number was determined by the optimal value of each index. The used indices included: KL index[4], CH index[5], Hartigan index[6], CCC index[7], Scott index[8], Marriot index[9], TrCovW index[10], TraceW index[10], Friedman index[11], Rubin index[11], Cindex[12], DB index[13], Silhouette index[14], Duda index[15], Pseudot2 index[15], Beale index[16], Ratkowsky index[17], Ball index[18], Ptbiserial index[19], Frey index[20], McClain index[21], Dunn index[22], Hubert index[23], SDindex[24], Dindex[25], and SDbw index[26]. More specifically, we set the cluster number k ranging from 2 to 8 and perform the clustering algorithm. Based on cluster structure identified given a specific k, the 21 indices are calculated. Each index measures how good the cluster structure is. For example, the maximum value of KL index indicates the optimal cluster structure given a specific cluster number. In this way, each of the 21 indices will suggest an optimal cluster number. Finally, the overall optimal cluster number can be determined by majority voting.

### D. Gaussian mixture model

Another clustering algorithm, Gaussian mixture model (GMM)[27], was used in sensitivity analysis to re-derive subphenotypes in the development cohort. Compared to agglomerative hierarchical clustering that is based on distance among data points, the GMM is a probabilistic model that assumes all the data points are generated from a mixture of a finite number of Gaussian distributions with unknown parameters. Typically, Akaike information criterion (AIC), Bayesian information criterion (BIC), and median probability of group membership are used to measure cluster structure of GMM to determine the optimal cluster number. In this analysis, we comprehensively considered these three measures to determine cluster number in sensitivity analysis.

### E. Outliers in sensitivity analysis

Outlier values were defined as values out of the range of $[\mu - 5\sigma, \mu + 5\sigma]$, where $\mu$ and $\sigma$ are the mean and standard deviation of the specific variable. Patients with outlier values were excluded for analysis.

### F. Leave-one-center-out analysis

To assess stability of the subphenotypes across the five medical centers, we further performed leave-one-center-out analysis. Specifically, within each loop of the leave-one-center-out procedure, we used a specific center as the within-loop validation cohort. The remaining four centers were combined as the within-loop development cohort which was used to derive clusters, i.e., subphenotypes, and train XGboost-based predictive model of subphenotypes. Then the predictive model was used to predict patients' cluster membership in the within-loop validation cohort. Similar to the consensus clustering[28], after all the five iterations, we generated a $N \times N$ cluster consensus matrix $M = [m_{ij}]$, where $N$ is the total number of patients and $m_{ij}$ is the ‘consensus value’ between patients $i$ and $j$, which is defined as the frequency that the two patients are assigned to the same cluster during the leave-one-center-out procedure. Typically, the consensus value ranges from 0 to 1, where a value of 0 means that the pair of patients never been grouped to the same cluster, while a value of 1 means that they always assigned to the same cluster during the leave-one-center-out procedure. Finally, the agglomerative hierarchical clustering was performed on the consensus matrix. Our goal is to confirm if we can still obtain cluster structure observed in our primary analysis under the leave-one-center-out procedure.

### G. Subphenotype visualization

#### G.1 Uniform manifold approximation and projection (UMAP) plots

UMAP is a novel dimension reduction technique that are widely used for data visualization[29]. Via a non-linear transformation, UMAP is able to project the high-dimensional data into a low-dimensional space, such that close data points in the original space will be close in the low-dimensional space, and vice versa. Compared to other dimension reduction methods, such as t-distributed stochastic neighbor embedding (t-SNE), UMAP is faster and better preserves the data's global structure. Therefore, we performed UMAP on the clinical variables to visualize patients in the 2-D space. Patients’ subphenotype membership were colored in the UMAP plots. In this study, UMAP plots were created by ‘umap-learn’ package in Python.

#### G.2 Chord diagram

Chord diagram plots were generated to visualize patterns of abnormal clinical variables and chronic comorbidities by subphenotypes, based on ‘Circlize’ package[30] in R.

*Subphenotype vs. abnormal clinical variables*. First of all, we grouped the clinical variables into the following categories:

- **Inflammatory markers**: C-reactive protein (CRP), erythrocyte sedimentation rate (ESR), interleukin 6 (IL-6), procalcitonin, bands, lactate dehydrogenase (LDH), lymphocyte count, neutrophil count, white blood cell count, albumin, and ferritin.
- **Hepatic markers**: albumin, ferritin, alanine aminotransferase (ALT), aspartate aminotransferase (AST), and bilirubin.
- **Cardiovascular markers**: creatine kinase (CK), lactate, troponin I, and troponin T.
- **Renal makers**: bicarbonate, blood urea nitrogen (BUN), creatinine, chloride, and sodium.
- **Hematologic markers**: d-dimer, hemoglobin, platelet count, prothrombin time, red blood cell distribution width (RDW), and glucose.

More specifically, for each subphenotype, if median of a clinical variable (e.g., CRP) is more abnormal than the median for the development cohort, we added a ribbon with one unit width between the subphenotype and the specific variable group (e.g., inflammatory markers). In this way, a broader ribbon between a subphenotype and a variable group means the subphenotype has more abnormal varaibles in this group compared to others.

*Subphenotypes vs. comorbidities*. We assessed nine chronic comorbidities, including hypertension, diabetes, coronary artery disease (CAD), heart failure, chronic obstructive pulmonary disease (COPD), asthma, cancer, obesity, and hyperlipidemia. In this context, each ribbon in the chord diagram indicates the normalized proportion of patients of a specific subphenotype with a specific comorbidity.

### eResults – Determination of optimal cluster number

#### A. Subphenotype derivation in development cohort

To derive subphenotype in development cohort, dendrogram showed that the 4-cluster model is the optimal fit of the agglomerative hierarchical clustering model (see eFigure 3 in Supplement). In addition, out of 21 indices in 'NbClust', 6 suggested 4 clusters, 5 suggested 2 clusters, 4 suggested 3 clusters, and 3 suggested 8 clusters. In conclusion, the optimal cluster number was 4.

#### B. Sensitivity analyses in development cohort

*Sensitivity to quality control and outliers*. A total of 140 patients with outlier values were excluded for re-deriving subphenotypes in development cohort. Performing agglomerative hierarchical clustering on the all 30 clinical variables of the remaining 8059 patients in development cohort, dendrogram showed that the 4-cluster model is the optimal fit (see eFigure 3 in Supplement). In addition, out of 21 indices in 'NbClust', 8 suggested 2 clusters, 6 suggested 4 clusters, and 4 suggested 3 clusters. Comprehensively considering the dendrogram and 'NbClust' indices, the optimal cluster number was 4.

*Sensitivity to clustering methods*. GMM was performed on the 23 clinical variables of all 8199 patients in development cohort. In GMM smaller AIC and BIC indicate better cluster structure in the data, while higher cluster membership probability of each data point indicates more robust result. By comprehensively considering the decreasing rates of AIC, BIC, and median of membership probability, we chose the optimal cluster number as 4 (see eTable 5 in Supplement).

#### C. Validation in internal validation cohort

To re-derive subphenotype in internal validation cohort, dendrogram also showed that the 4-cluster model is the optimal fit of the agglomerative hierarchical clustering model (see eFigure 7 in Supplement). In addition, out of 21 indices in 'NbClust', 7 suggested 2 clusters, 7 suggested 3 clusters, and 6 suggested 4 clusters. Comprehensively considering the dendrogram and 'NbClust' indices, the optimal cluster number was 4.

#### D. Derivation of SDoH strata in development cohort

In the development cohort, 7862 patients who had SDoH data were used to derive SDoH strata. Dendrogram showed that the 3-cluster model is the optimal fit of the agglomerative hierarchical clustering model (see eFigure 13 in Supplement). In addition, out of 21 indices in 'NbClust', 9 suggested 3 clusters, 6 suggested 2 clusters, 3 suggested 4 clusters, and 3 suggested 8 clusters. Therefore, the optimal cluster number was 4.

**eFigure 1. Inclusion-exclusion criteria**

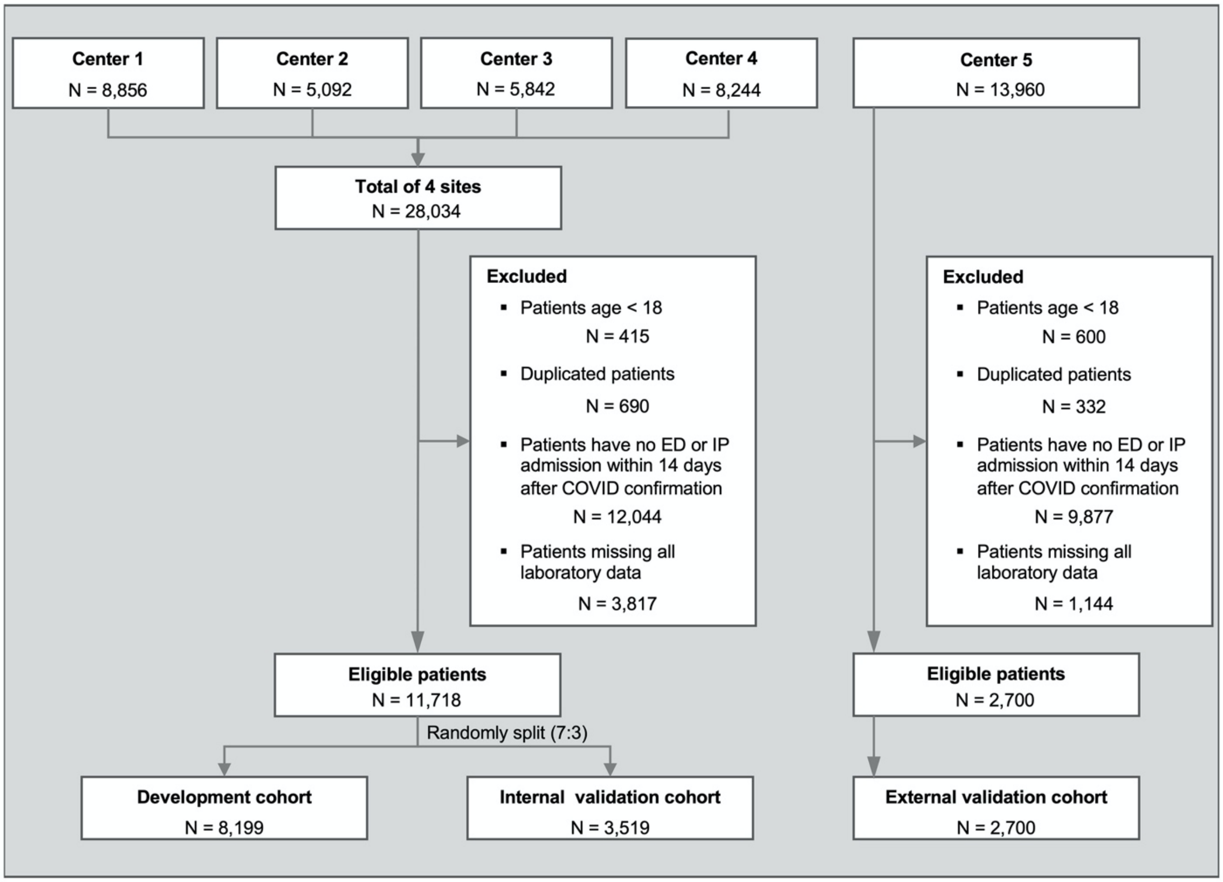

### eFigure 2. Method for extracting clinical data of patients

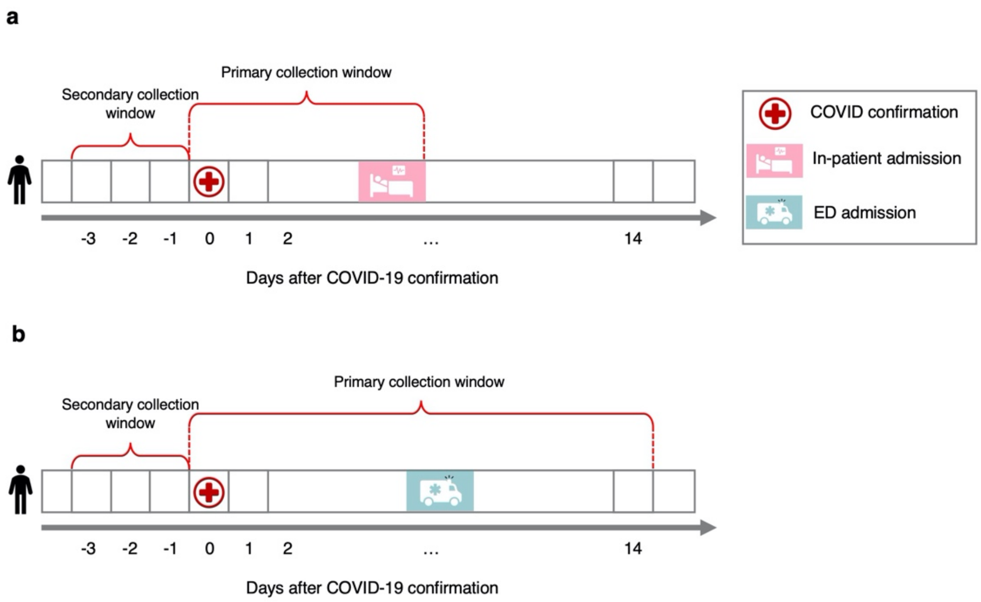

The primary collection window was defined as **(a)** time period from COVID-19 confirmation to the first in-patient encounter, if the patient has in-patient admission within 14 days after confirmation; or **(b)** 14 days after COVID-19 confirmation otherwise. We extracted the first value of each clinical variable within the primary collection window for each patient. If there was no record in the primary collection window, we extracted the last value within 3 days before confirmation (secondary collection window).

Abbreviations: ED = emergency department

**eFigure 3. Dendrogram for the subphenotypes in the development cohort**

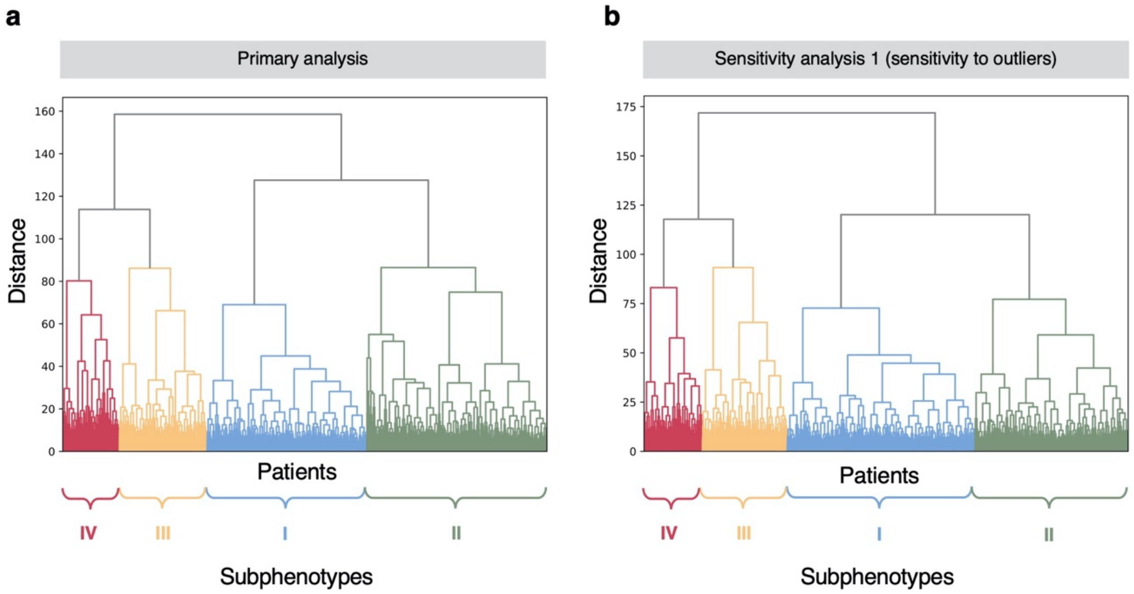

### eFigure 4. UMAP-based visualization of the subphenotypes in the development cohort

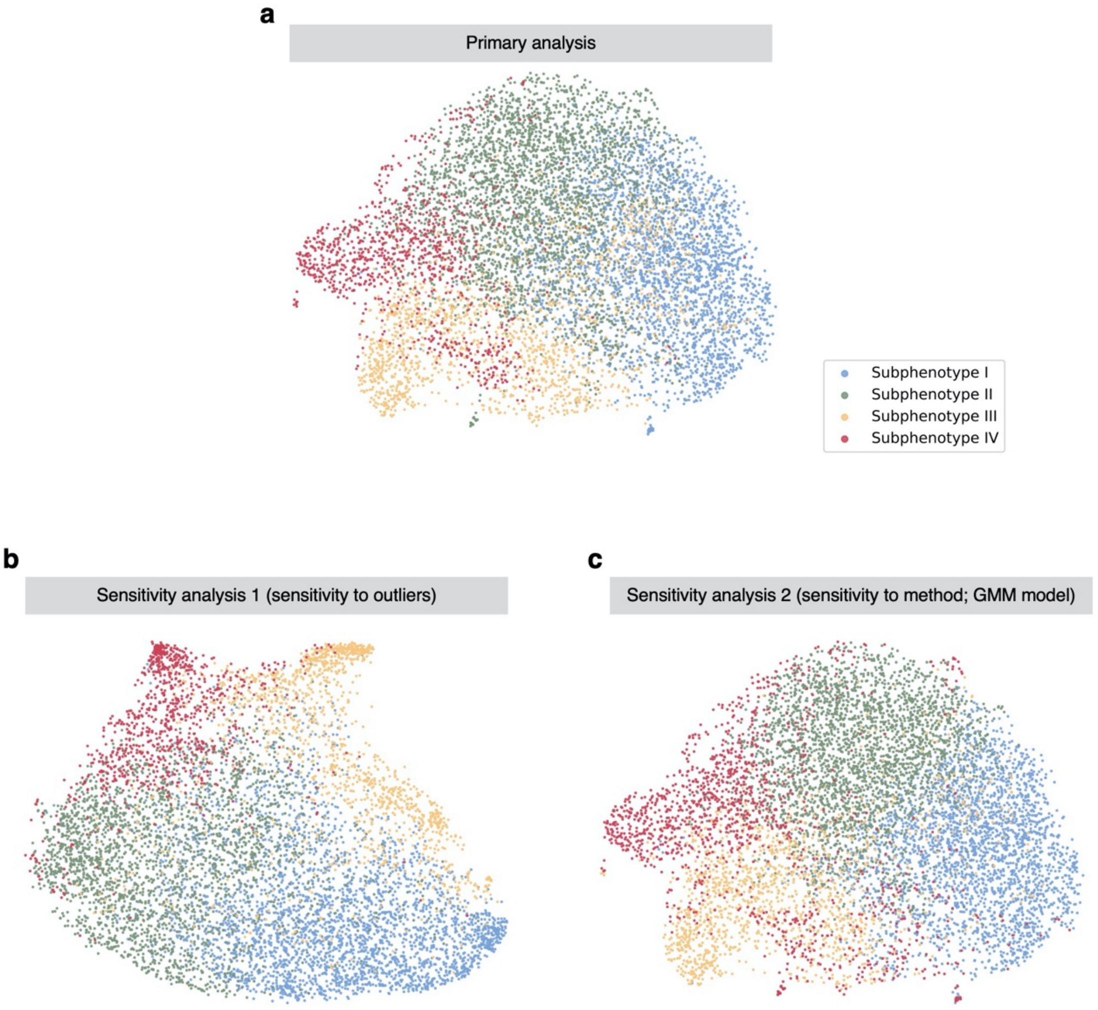

Abbreviations: UMAP = uniform manifold approximation and projection for dimension reduction.

**eFigure 5. ICU admission and intubation outcomes by subphenotype in the development cohort**

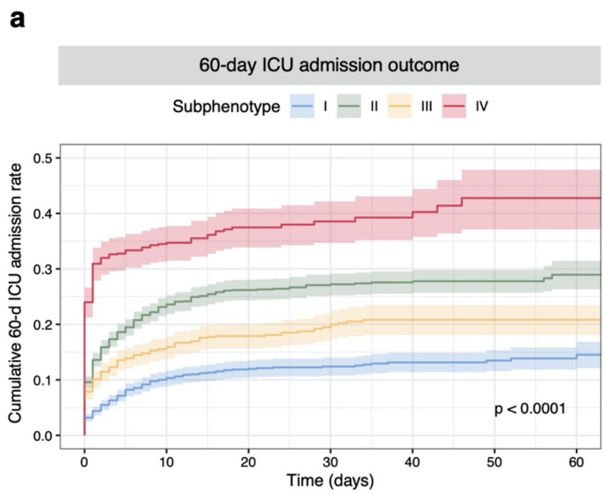

**eFigure 6. Confusion matrices for comparing the subphenotypes derived by primary analysis and sensitivity analyses in the development cohort**

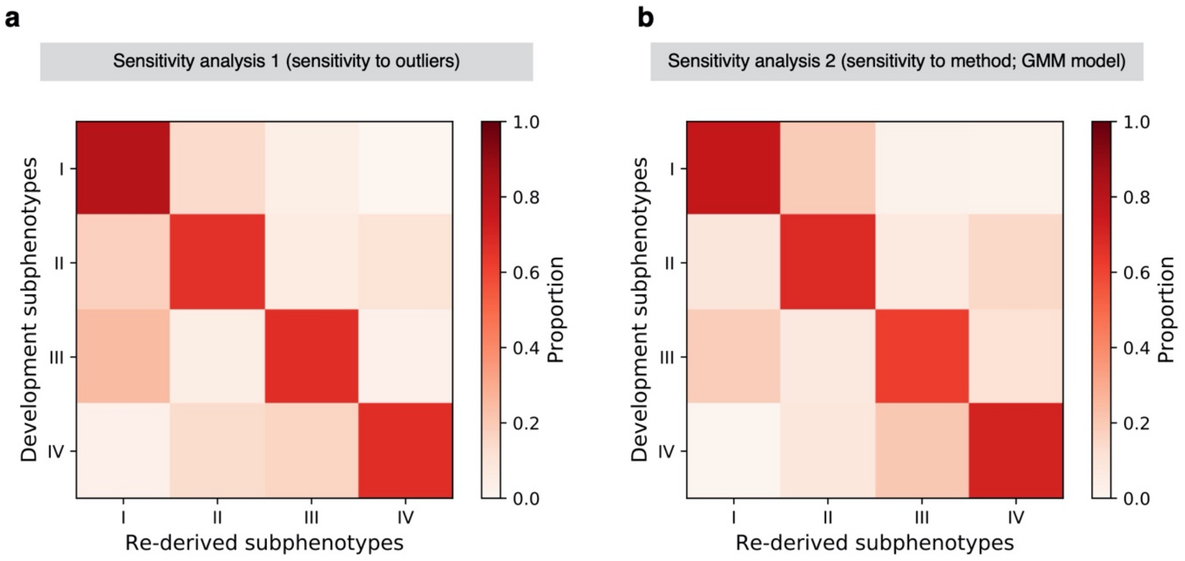

**eFigure 7. Dendrogram and UMAP-based visualization for the subphenotypes in the internal validation cohort**

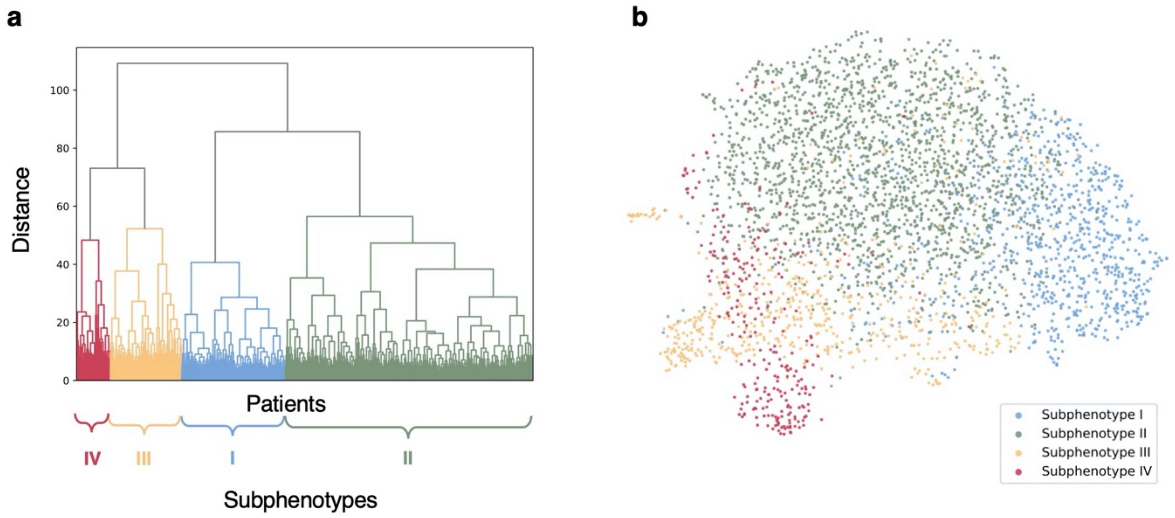

Abbreviations: UMAP = uniform manifold approximation and projection for dimension reduction.

**eFigure 8. Chord diagrams showing abnormal clinical variables and comorbidities by subphenotype in the internal validation cohort**

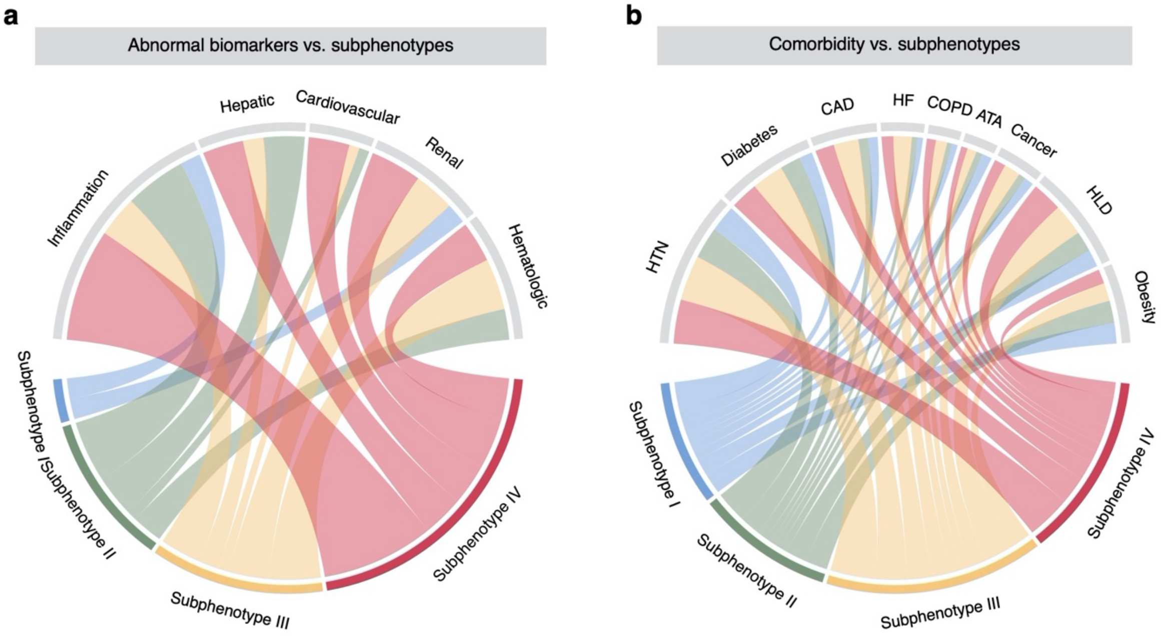

Abbreviations: ATA = asthma; CAD = coronary artery disease; COPD = chronic obstructive pulmonary disease; HF = heart failure; HLD = hyperlipidemia; HTN = hypertension.

**eFigure 9. Receiver operating characteristic (ROC) curves of the subphenotype prediction model in the development cohort**

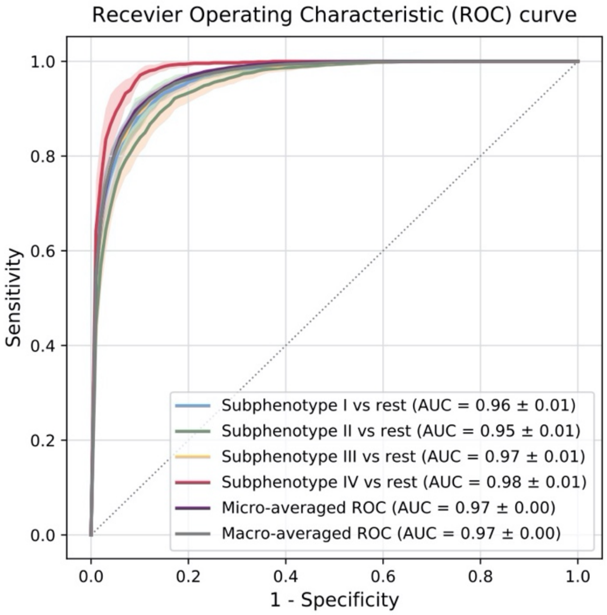

A macro-average typically computes the metric independently for each class and then takes the average (hence treating all classes equally), whereas a micro-average tries to aggregate the contributions of all classes to compute the average metric.

Abbreviations: AUC = area under the receiver operating characteristic curve.

**eFigure 10. SHAP value-based predictor contribution visualization of the subphenotype prediction model**

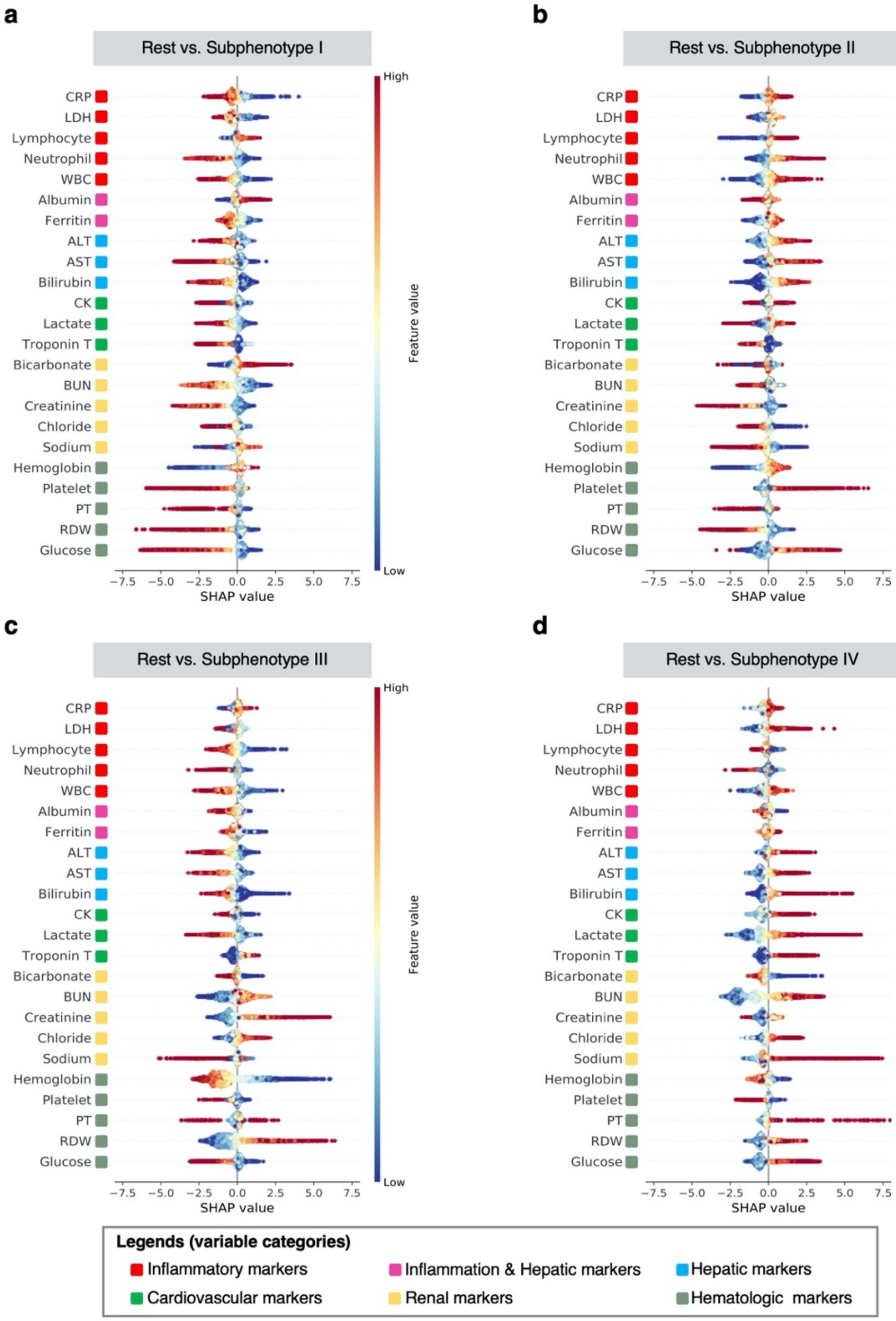

Abbreviations: ALT = alanine aminotransferase; AST = aspartate aminotransferase; BUN = blood urea nitrogen; CK = creatine kinase; CRP = C-reactive protein; ESR = Erythrocyte sedimentation rate; LDH = Lactate dehydrogenase; PT = prothrombin time; RDW = red blood cell distribution width; WBC = white blood cell count

**eFigure 11. UMAP-based visualization for the subphenotypes in the external validation cohort**

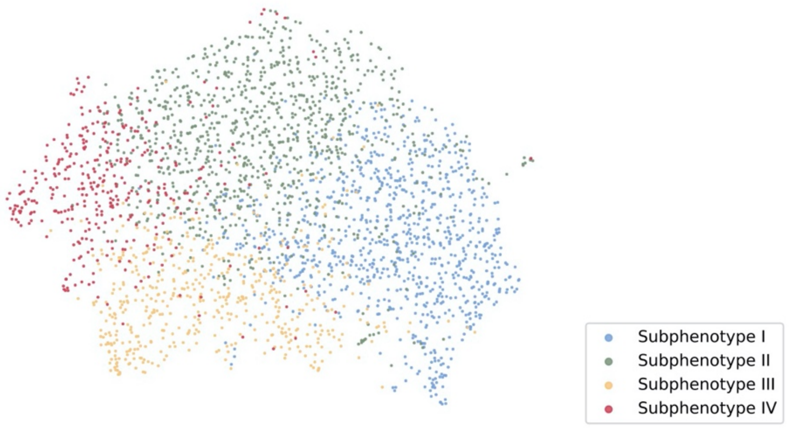

Abbreviations: UMAP = uniform manifold approximation and projection for dimension reduction.

**eFigure 12. Chord diagrams showing abnormal clinical variables and comorbidities by subphenotype in the external validation cohort**

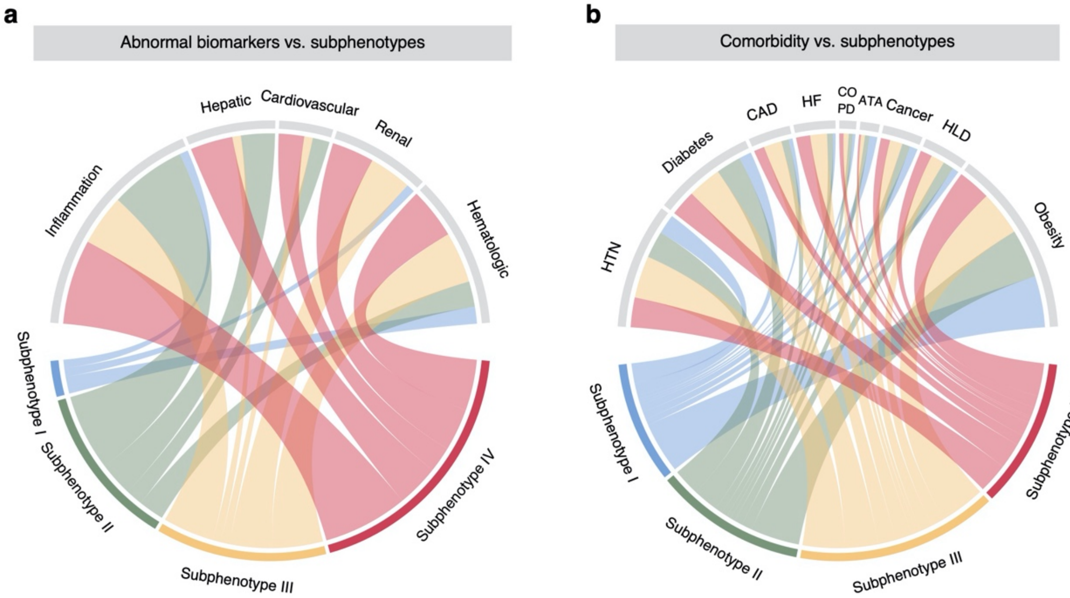

Abbreviations: ATA = asthma; CAD = coronary artery disease; COPD = chronic obstructive pulmonary disease; HF = heart failure; HLD = hyperlipidemia; HTN = hypertension.

**eFigure 13. Consensus matrix showing clusters obtained in leave-one-center-out cross-validation.** Density of color denotes 'consensus value' between each pair of patients, calculated based on the frequency that these two patients fall into same cluster in the leave-one-center-out procedure. A consensus value of 0 means that the pair of patients never been grouped into the same cluster, while a value of 1 means that they always assigned to the same cluster during the leave-one-center-out procedure. By agglomerative hierarchical clustering, we identified clear four-cluster structure, demonstrating stability of the clusters (i.e., subphenotypes) across the five studied centers.

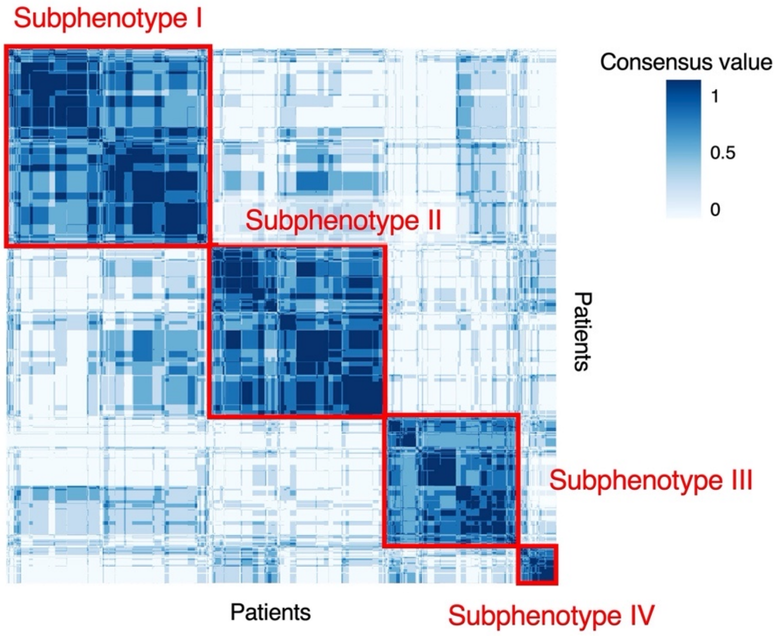

**eFigure 14. Dendrogram for SDoH strata identification in the development cohort**

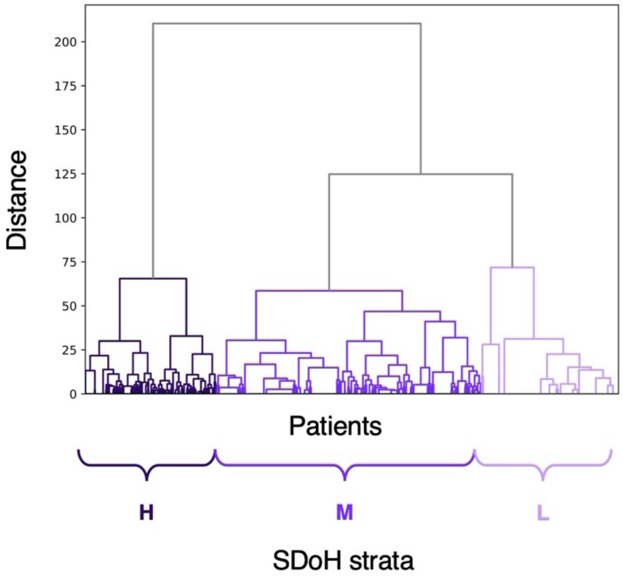

Abbreviations: SDoH = social determinants of health.

**eTable 1. Characteristics of included patients in the five medical centers.**

| Variable | Medical centers | | | | |
|---|---|---|---|---|---|
| | Center 1 | Center 2 | Center 3 | Center 4 | Center 5 |
| No. of patients | 4348 | 1215 | 314 | 5841 | 2700 |
| **Demographics** | | | | | |
| Age, y, Median [IQR] | 63.60 [49.96 - 75.18] | 63.08 [49.15 - 76.23] | 60.00 [45.55 - 71.20] | 63.85 [51.59 - 75.13] | 65.58 [51.08 - 77.39] |
| Sex female, N (%) | 1878 (43.2%) | 565 (46.5%) | 157 (50.0%) | 2772 (47.5%) | 1305 (48.33) |
| Race, N (%) | | | | | |
| White | 1874 (43.1%) | 366 (30.1%) | 65 (20.7%) | 569 (9.7%) | 675 (25.00) |
| Black | 697 (16.0%) | 169 (13.9%) | 72 (22.9%) | 2132 (36.5%) | 545 (20.19) |
| Asian | 279 (6.4%) | 162 (13.3%) | 9 (2.9%) | 152 (2.6%) | 28 (1.04) |
| Other/unknown | 1498 (34.5%) | 518 (42.6%) | 168 (53.5%) | 2988 (51.2%) | 1452 (53.78) |
| **Patient care** | | | | | |
| Hospitalized patients[a], N (%) | 641 (14.7%) | 136 (11.2%) | 198 (63.1%) | 1016 (17.4%) | 363 (13.4%) |
| Patients presented to ED without hospitalization[b], N (%) | 3707 (85.3%) | 1079 (88.8%) | 116 (36.9%) | 4825 (82.6%) | 2337 (86.6%) |

Abbreviations: ED = emergency department; IQR = Interquartile range.

[a] Patients who hospitalized within 14 days after COVID-19 confirmation.

[b] Patients who only had ED admission within 14 days after COVID-19 confirmation.

### eTable 2. Presenting value distribution of candidate clinical variables for cluster analysis

| Variable | Development cohort | Internal validation cohort | External validation cohort |
|---|---|---|---|
| Albumin, g/dL, Median [IQR] | 3.70 [3.30 - 4.10] | 3.70 [3.30 - 4.10] | 3.80 [3.40 - 4.10] |
| Alanine aminotransferase, U/L, Median [IQR] | 29.00 [19.00 - 48.00] | 30.00 [19.00 - 49.00] | 39.00 [26.00 - 65.00] |
| Aspartate aminotransferase, U/L, Median [IQR] | 39.00 [26.00 - 63.00] | 40.00 [27.00 - 64.00] | 27.00 [17.00 - 47.00] |
| Bands, %, Median [IQR] | 2.00 [0.00 - 5.00] | 2.00 [1.00 - 6.00] | 3.00 [2.00 - 5.00] |
| Bicarbonate, mmol/L, Median [IQR] | 23.00 [21.00 - 26.00] | 23.00 [21.00 - 26.00] | 23.00 [20.00 - 25.00] |
| Bilirubin, mg/dL, Median [IQR] | 0.30 [0.20 - 0.60] | 0.30 [0.20 - 0.60] | 0.30 [0.20 - 0.50] |
| BUN, mg/dL, Median [IQR] | 17.00 [11.00 - 31.00] | 17.00 [11.00 - 29.00] | 18.00 [12.00 - 33.00] |
| Creatine kinase, U/L, Median [IQR] | 154.00 [78.00 - 359.00] | 151.00 [80.50 - 335.50] | 154.50 [79.00 - 347.75] |
| Creatinine, mg/dL, Median [IQR] | 1.00 [0.80 - 1.50] | 1.00 [0.80 - 1.50] | 1.10 [0.80 - 1.60] |
| Chloride, mmol/L, Median [IQR] | 100.00 [97.00 - 104.00] | 100.00 [97.00 - 104.00] | 100.00 [96.00 - 103.00] |
| C-reactive protein, mg/L, Median [IQR] | 9.40 [3.70 – 16.80] | 9.63 [4.39 – 16.42] | 11.30 [5.15 – 20.15] |
| D-dimer, ng/mL, Median [IQR] | 1360.00 [620.00 - 3370.00] | 1330.00 [585.25 - 3092.50] | 1400.00 [700.00 - 3100.00] |
| ESR, mm/hr, Median [IQR] | 69.00 [42.00 - 97.00] | 64.00 [41.00 - 93.00] | 72.00 [50.00 - 97.00] |
| Ferritin, ng/mL, Median [IQR] | 645.00 [295.90 - 1347.00] | 699.50 [297.00 - 1435.00] | 672.65 [333.03 - 1276.00] |
| Glucose, mg/dL, Median [IQR] | 121.00 [101.00 - 165.00] | 122.00 [101.50 - 164.00] | 125.00 [104.00 - 176.00] |
| Hemoglobin, g/dL, Median [IQR] | 13.10 [11.50 - 14.60] | 13.30 [11.70 - 14.70] | 13.00 [11.40 - 14.30] |
| IL-6, pg/mL, Median [IQR] | 19.00 [10.00 - 42.00] | 16.50 [9.00 - 35.25] | 45.20 [11.15 - 108.40] |
| Lactate, mmol/L, Median [IQR] | 1.90 [1.40 - 2.60] | 1.90 [1.30 - 2.60] | 1.60 [1.20 - 2.30] |
| LDH, U/L, Median [IQR] | 377.00 [280.00 - 525.00] | 382.00 [285.00 - 527.00] | 419.00 [310.50 - 599.50] |
| Lymphocyte count, $\times 10^3$/uL, Median [IQR] | 1.00 [0.70 - 1.43] | 1.00 [0.70 - 1.40] | 1.00 [0.70 - 1.50] |
| Neutrophil count, $\times 10^3$/uL, Median [IQR] | 5.30 [3.70 - 7.90] | 5.30 [3.70 - 7.70] | 5.40 [3.70 - 8.10] |
| Oxygen saturation, %, Median [IQR] | 69.00 [50.00 - 85.00] | 72.00 [48.50 - 90.00] | 97.10 [92.00 - 98.70] |
| Prothrombin time, s, Median [IQR] | 13.30 [12.20 - 14.60] | 13.20 [12.20 - 14.40] | 14.20 [13.40 - 15.30] |
| Procalcitonin, ng/mL, Median [IQR] | 0.20 [0.10 - 0.60] | 0.20 [0.10 - 0.50] | 0.20 [0.10 - 0.60] |
| Platelet count, $\times 10^3$/uL, Median [IQR] | 211.00 [162.00 - 277.00] | 210.00 [163.00 - 274.00] | 203.00 [157.00 - 262.50] |
| Red blood cell distribution width, %, Median [IQR] | 13.80 [12.90 - 15.00] | 13.70 [13.00 - 14.90] | 13.90 [13.00 - 15.00] |
| Sodium, mmol/L, Median [IQR] | 137.00 [134.00 - 140.00] | 137.00 [134.00 - 140.00] | 138.00 [135.00 - 141.00] |
| Troponin I, ng/mL, Median [IQR] | 0.10 [0.06 - 0.30] | 0.10 [0.07 - 0.30] | 0.00 [0.00 - 0.10] |

| | | | |
|---|---|---|---|
| Troponin T, ng/mL, Median [IQR] | 0.01 [0.01 - 0.03] | 0.01 [0.01 - 0.02] | 0.02 [0.01 - 0.04] |
| White blood cell count, $\times 10^3$/uL, Median [IQR] | 7.20 [5.30 - 9.90] | 7.10 [5.30 - 9.60] | 7.30 [5.40 - 10.20] |

Abbreviations: BUN = blood urea nitrogen; ESR = Erythrocyte sedimentation rate; IL-6 = Interleukin 6; IQR = Interquartile range; LDH = Lactate dehydrogenase.

**eTable 3. Missing values (n (%)) across cohorts for candidate clinical variables at presentation**

| Variable | Development cohort | Internal validation cohort | External validation cohort |
|---|---|---|---|
| No. of patients | 8,199 | 3,519 | 2,700 |
| Albumin | 915 (11.2%) | 404 (11.5%) | 476 (17.6%) |
| Alanine aminotransferase | 961 (11.7%) | 422 (12.0%) | 474 (17.6%) |
| Aspartate aminotransferase | 1383 (16.9%) | 613 (17.4%) | 467 (17.3%) |
| Bands | 6435 (78.5%) | 2805 (79.7%) | 2467 (91.4%) |
| Bicarbonate | 254 (3.1%) | 138 (3.9%) | 188 (7.0%) |
| Bilirubin | 989 (12.1%) | 442 (12.6%) | 474 (17.6%) |
| BUN | 209 (2.5%) | 115 (3.3%) | 195 (7.2%) |
| Creatine kinase | 3958 (48.3%) | 1752 (49.8%) | 1394 (51.6%) |
| Creatinine | 202 (2.5%) | 111 (3.2%) | 187 (6.9%) |
| Chloride | 364 (4.4%) | 195 (5.5%) | 187 (6.9%) |
| C-reactive protein | 3697 (45.1%) | 1617 (46.0%) | 1109 (41.1%) |
| D-dimer | 6119 (74.6%) | 2651 (75.3%) | 1869 (69.2%) |
| ESR | 6834 (83.4%) | 2907 (82.6%) | 1465 (54.3%) |
| Ferritin | 4120 (50.3%) | 1814 (51.5%) | 1240 (45.9%) |
| Glucose | 199 (2.4%) | 92 (2.6%) | 143 (5.3%) |
| Hemoglobin | 52 (0.6%) | 28 (0.8%) | 103 (3.8%) |
| IL-6 | 7685 (93.7%) | 3287 (93.4%) | 2380 (88.1%) |
| Lactate | 3444 (42.0%) | 1498 (42.6%) | 1129 (41.8%) |
| LDH | 5422 (66.1%) | 2326 (66.1%) | 1197 (44.3%) |
| Lymphocyte count | 659 (8.0%) | 277 (7.9%) | 275 (10.2%) |
| Neutrophil count | 679 (8.3%) | 287 (8.2%) | 272 (10.1%) |
| Oxygen saturation | 7604 (92.7%) | 3236 (92.0%) | 2465 (91.3%) |
| Prothrombin time | 5552 (67.7%) | 2397 (68.1%) | 1105 (40.9%) |
| Procalcitonin | 5973 (72.9%) | 2568 (73.0%) | 1035 (38.3%) |
| Platelet count | 88 (1.1%) | 47 (1.3%) | 129 (4.8%) |
| Red blood cell distribution width | 82 (1.0%) | 41 (1.2%) | 119 (4.4%) |
| Sodium | 198 (2.4%) | 107 (3.0%) | 176 (6.5%) |
| Troponin T | 4939 (60.2%) | 2157 (61.3%) | 831 (30.8%) |
| Troponin I | 7236 (88.3%) | 3114 (88.5%) | 2597 (96.2%) |
| White blood cell count | 78 (1.0%) | 40 (1.1%) | 119 (4.4%) |

Abbreviations: BUN = blood urea nitrogen; ESR = Erythrocyte sedimentation rate; IL-6 = Interleukin 6; IQR = Interquartile range; LDH = Lactate dehydrogenase.

### eTable 4. Usage of the candidate clinical variables in analysis

| Variable | **Usage of the clinical variable in analysis**<br>D: Used for subphenotype derivation (development cohort).<br>S: Used for sensitivity analysis.<br>R: Used for re-derivation of subphenotypes (validation cohort).<br>P: Used for building prediction model of subphenotypes | Logarithmic transform? |
|---|---|---|
| Albumin | D, S, R, P | |
| Alanine aminotransferase | D, S, R, P | yes |
| Aspartate aminotransferase | D, S, R, P | yes |
| Bands | S | yes |
| Bicarbonate | D, S, R, P | |
| Bilirubin | D, S, R, P | yes |
| BUN | D, S, R, P | yes |
| Creatine kinase | D, S, R, P | yes |
| Creatinine | D, S, R, P | yes |
| Chloride | D, S, R, P | yes |
| C-reactive protein | D, S, R, P | yes |
| D-dimer | S | yes |
| ESR | S | |
| Ferritin | D, S, R, P | yes |
| Glucose | D, S, R, P | yes |
| Hemoglobin | D, S, R, P | yes |
| IL-6 | S | yes |
| Lactate | D, S, R, P | yes |
| LDH | D, S, R, P | yes |
| Lymphocyte count | D, S, R, P | yes |
| Neutrophil count | D, S, R, P | |
| Oxygen saturation | S | |
| Prothrombin time | D, S, R, P | yes |
| Procalcitonin | S | yes |
| Platelet count | D, S, R, P | |
| Red blood cell distribution width | D, S, R, P | |
| Sodium | D, S, R, P | yes |
| Troponin T | D, S, R, P | yes |
| Troponin I | S | yes |
| White blood cell count | D, S, R, P | yes |

Abbreviations: BUN = blood urea nitrogen; ESR = Erythrocyte sedimentation rate; IL-6 = Interleukin 6; IQR = Interquartile range; LDH = Lactate dehydrogenase.

### eTable 5. Statistical output from Gaussian mixture model for sensitivity analysis 2 in development cohort

| Cluster number | BIC | AIC | Membership probability, median [IQR] |
|---|---|---|---|
| 2 | 469836.062 | 469492.486 | 99.99 [99.44 - 100.00] |
| 3 | 459066.27 | 458547.399 | 99.74 [94.16 - 100.00] |
| 4 | 454755.227 | 454061.062 | 99.11 [88.76 - 99.97] |
| 5 | 449328.294 | 448458.834 | 98.22 [85.53 - 99.90] |
| 6 | 446186.472 | 445141.719 | 97.29 [82.43 - 99.83] |
| 7 | 443065.314 | 441845.266 | 97.43 [82.84 - 99.84] |
| 8 | 440346.512 | 438951.17 | 97.30 [82.37 - 99.81] |

Abbreviations: IQR = Interquartile range

**eTable 6. Characteristics of the subphenotypes in sensitivity analysis 1 (quality control and outliers) in the development cohort.**

| Variable | Total | Subphenotype I | Subphenotype II | Subphenotype III | Subphenotype IV | P-value[a] | P-value (age and sex adjusted)[b] |
|---|---|---|---|---|---|---|---|
| No. of patients (%) | 8059 (100)[c] | 3125 (38.78) | 2549 (31.63) | 1413 (17.53) | 972 (12.06) | - | - |
| **Demographics** | | | | | | | |
| Age, y, Median (IQR) | 63.53 (50.54 - 75.17) | 60.47 (46.11 - 73.36) | 61.10 (49.95 - 71.13) | 67.63 (55.55 - 79.35) | 72.92 (62.46 - 82.62) | < 0.001 | - |
| Sex female, N (%) | 3723 (46.20) | 1723 (55.14) | 908 (35.62) | 695 (49.19) | 397 (40.84) | < 0.001 | - |
| Race, N (%) | | | | | | | |
| White | 2001 (24.83) | 856 (27.39) | 603 (23.66) | 331 (23.43) | 211 (21.71) | < 0.001 | - |
| Black | 2111 (26.19) | 806 (25.79) | 520 (20.40) | 472 (33.40) | 313 (33.20) | | |
| Asian | 399 (4.95) | 138 (4.42) | 174 (6.83) | 53 (3.75) | 34 (3.5) | | |
| Other/unknown | 3548 (44.02) | 1325 (42.40) | 1252 (49.12) | 557 (39.42) | 414 (42.60) | | |
| **Inflammatory markers** | | | | | | | |
| C-reactive protein, mg/L, Median (IQR) | 9.35 (3.78 - 16.77) | 4.60 (1.40 - 9.35) | 13.20 (7.44 - 19.56) | 8.60 (3.20 - 15.49) | 16.24 (7.57 - 25.10) | < 0.001 | < 0.001 |
| ESR, mm/hr, Median (IQR) | 69.00 (42.00 - 97.00) | 51.00 (33.00 - 76.00) | 76.00 (51.00 - 99.00) | 76.00 (47.00 - 106.25) | 86.00 (50.50 - 108.00) | < 0.001 | < 0.001 |
| IL-6, pg/mL, Median (IQR) | 18.00 (10.00 - 42.00) | 13.00 (8.00 - 26.25) | 17.50 (10.00 - 41.00) | 20.00 (11.00 - 47.00) | 37.00 (17.00 - 73.50) | < 0.001 | < 0.001 |
| Procalcitonin, ng/mL, Median (IQR) | 0.20 (0.10 - 0.51) | 0.10 (0.10 - 0.20) | 0.20 (0.10 - 0.50) | 0.40 (0.14 - 1.12) | 0.64 (0.30 - 2.08) | < 0.001 | < 0.001 |
| Bands, %, Median (IQR) | 2.00 (0.00 - 5.00) | 3.00 (0.00 - 6.00) | 2.00 (1.00 - 5.00) | 2.00 (0.00 - 5.00) | 2.00 (0.00 - 6.00) | 0.32 | 0.09 |
| LDH, U/L, Median (IQR) | 376.00 (279.00 - 521.50) | 301.00 (237.00 - 388.50) | 433.00 (345.00 - 574.00) | 341.00 (256.00 - 477.00) | 567.00 (420.00 - 789.00) | < 0.001 | < 0.001 |
| Lymphocyte count, $\times 10^3$/uL, Median (IQR) | 1.00 (0.70 - 1.40) | 1.10 (0.80 - 1.60) | 0.90 (0.60 - 1.30) | 0.90 (0.60 - 1.30) | 1.00 (0.70 - 1.50) | < 0.001 | < 0.001 |
| Neutrophil count, $\times 10^3$/uL, Median (IQR) | 5.30 (3.60 - 7.80) | 4.20 (3.00 - 5.60) | 6.10 (4.30 - 8.50) | 5.30 (3.70 - 7.40) | 10.41 (7.50 - 13.78) | < 0.001 | < 0.001 |
| White blood cell count, $\times 10^3$/uL, Median (IQR) | 7.20 (5.30 - 9.80) | 6.00 (4.70 - 7.70) | 7.90 (6.00 - 10.30) | 7.00 (5.20 - 9.50) | 12.70 (9.50 - 16.50) | < 0.001 | < 0.001 |
| **Inflammation & Hepatic markers** | | | | | | | |
| Albumin, g/dL, Median (IQR) | 3.70 (3.30 - 4.10) | 3.90 (3.60 - 4.30) | 3.70 (3.30 - 4.00) | 3.40 (2.90 - 3.90) | 3.40 (2.90 - 3.73) | < 0.001 | < 0.001 |
| Ferritin, ng/mL, Median (IQR) | 643.00 (296.00 - 1339.60) | 351.55 (173.97 - 642.35) | 958.00 (503.80 - 1666.70) | 621.20 (212.00 - 1506.10) | 1026.10 (519.00 - 1987.25) | < 0.001 | < 0.001 |
| **Hepatic markers** | | | | | | | |
| Alanine aminotransferase, U/L, Median (IQR) | 29.00 (19.00 - 48.00) | 24.00 (17.00 - 34.00) | 45.00 (28.00 - 73.00) | 20.00 (14.00 - 32.00) | 39.00 (23.00 - 67.00) | < 0.001 | < 0.001 |
| Aspartate aminotransferase, U/L, Median (IQR) | 39.00 (26.00 - 63.00) | 31.00 (23.00 - 41.00) | 57.00 (39.00 - 84.00) | 32.00 (21.00 - 50.00) | 62.00 (36.00 - 113.00) | < 0.001 | < 0.001 |
| Bilirubin, mg/dL, Median (IQR) | 0.30 (0.20 - 0.60) | 0.30 (0.20 - 0.50) | 0.40 (0.20 - 0.60) | 0.30 (0.20 - 0.50) | 0.40 (0.20 - 0.60) | < 0.001 | < 0.001 |
| **Cardiovascular markers** | | | | | | | |
| Creatine kinase, U/L, Median (IQR) | 153.00 (78.00 - 353.00) | 123.00 (75.00 - 217.25) | 190.00 (96.00 - 441.50) | 117.00 (57.50 - 283.50) | 305.00 (110.00 - 1041.00) | < 0.001 | < 0.001 |
| Lactate, mmol/L, Median (IQR) | 1.80 (1.38 - 2.60) | 1.60 (1.20 - 2.12) | 1.90 (1.50 - 2.50) | 1.70 (1.30 - 2.30) | 3.10 (2.20 - 4.70) | < 0.001 | < 0.001 |
| Troponin I, ng/mL, Median (IQR) | 0.10 (0.06 - 0.30) | 0.10 (0.01 - 0.10) | 0.10 (0.04 - 0.28) | 0.10 (0.10 - 0.30) | 0.20 (0.10 - 0.50) | < 0.001 | 0.02 |
| Troponin T, ng/mL, Median (IQR) | 0.01 (0.01 - 0.03) | 0.01 (0.01 - 0.01) | 0.01 (0.01 - 0.01) | 0.04 (0.01 - 0.12) | 0.04 (0.01 - 0.12) | < 0.001 | < 0.001 |
| **Renal markers** | | | | | | | |
| Bicarbonate, mmol/L, Median (IQR) | 23.00 (21.00 - 26.00) | 24.00 (22.00 - 26.00) | 23.00 (21.00 - 26.00) | 23.00 (19.00 - 25.10) | 20.00 (17.00 - 23.00) | < 0.001 | < 0.001 |

| | | | | | | | |
|---|---|---|---|---|---|---|---|
| BUN, mg/dL, Median (IQR) | 17.00 (11.00 - 30.00) | 14.00 (10.00 - 19.00) | 15.00 (11.00 - 22.00) | 30.00 (16.00 - 56.00) | 47.00 (28.00 - 79.00) | < 0.001 | < 0.001 |
| Creatinine, mg/dL, Median (IQR) | 1.00 (0.80 - 1.50) | 0.90 (0.74 - 1.12) | 0.97 (0.80 - 1.20) | 1.70 (0.90 - 4.80) | 1.85 (1.20 - 3.08) | < 0.001 | < 0.001 |
| Chloride, mmol/L, Median (IQR) | 100.00 (97.00 - 104.00) | 101.00 (98.00 - 104.00) | 98.00 (95.00 - 101.00) | 101.00 (96.00 - 105.00) | 104.00 (98.00 - 112.00) | < 0.001 | < 0.001 |
| Sodium, mmol/L, Median (IQR) | 137.00 (134.00 - 140.00) | 138.00 (136.00 - 141.00) | 135.00 (132.00 - 138.00) | 137.00 (134.00 - 140.00) | 141.00 (135.00 - 152.00) | < 0.001 | < 0.001 |
| **Hematologic markers** | | | | | | | |
| D-dimer, ng/mL, Median (IQR) | 1345.00 (611.50 - 3332.50) | 706.00 (380.00 - 1470.00) | 1080.00 (590.00 - 2185.00) | 2030.00 (990.00 - 3680.00) | 4640.00 (2250.00 - 13960.00) | < 0.001 | < 0.001 |
| Hemoglobin, g/dL, Median (IQR) | 13.20 (11.60 - 14.60) | 13.40 (12.20 - 14.70) | 13.80 (12.60 - 15.10) | 10.60 (8.70 - 12.10) | 13.20 (11.45 - 15.20) | < 0.001 | < 0.001 |
| Platelet count, $\times 10^3$/uL, Median (IQR) | 211.00 (163.00 - 276.00) | 201.00 (161.00 - 253.00) | 212.00 (168.00 - 271.00) | 211.00 (148.00 - 309.00) | 253.00 (177.00 - 344.00) | < 0.001 | < 0.001 |
| Prothrombin time, s, Median (IQR) | 13.20 (12.20 - 14.50) | 12.80 (12.00 - 13.80) | 13.40 (12.40 - 14.50) | 13.30 (12.00 - 15.60) | 14.40 (13.00 - 16.95) | < 0.001 | < 0.001 |
| Red blood cell distribution width, %, Median (IQR) | 13.80 (12.90 - 15.00) | 13.50 (12.80 - 14.40) | 13.20 (12.60 - 14.10) | 15.80 (14.20 - 17.90) | 14.70 (13.70 - 16.10) | < 0.001 | < 0.001 |
| Glucose, mg/dL, Median (IQR) | 121.00 (101.00 - 165.00) | 110.00 (96.00 - 131.00) | 134.00 (110.00 - 210.00) | 117.00 (96.00 - 154.00) | 158.00 (120.00 - 239.00) | < 0.001 | < 0.001 |
| **Other markers** | | | | | | | |
| Oxygen saturation, %, Median (IQR) | 69.00 (50.00 - 85.00) | 67.00 (49.00 - 85.00) | 69.00 (52.00 - 85.00) | 70.50 (48.00 - 80.75) | 76.00 (53.25 - 91.65) | 0.31 | 0.41 |
| BMI, kg/m$^2$, Median (IQR) | 28.00 (25.00 - 33.00) | 29.00 (25.00 - 34.00) | 29.00 (25.00 - 33.00) | 27.00 (23.00 - 32.00) | 26.00 (23.00 - 31.00) | < 0.001 | 0.62 |
| **Comorbidity, (missing=579), N (%)** | | | | | | | |
| Hypertension | 4664 (62.35) | 1571 (54.34) | 1373 (58.25) | 1015 (76.89) | 705 (77.30) | < 0.001 | - |
| Diabetes | 3057 (40.87) | 867 (29.99) | 1018 (43.19) | 682 (51.67) | 490 (53.73) | < 0.001 | - |
| Coronary artery disease | 1719 (22.98) | 507 (17.54) | 420 (17.82) | 490 (37.12) | 302 (33.11) | < 0.001 | - |
| Heart failure | 1099 (14.69) | 290 (10.03) | 197 (8.36) | 418 (31.67) | 194 (21.27) | < 0.001 | - |
| COPD | 945 (12.63) | 345 (11.93) | 190 (8.06) | 259 (19.62) | 151 (16.56) | < 0.001 | - |
| Asthma | 1069 (14.29) | 441 (15.25) | 318 (13.49) | 217 (16.44) | 93 (10.20) | < 0.001 | - |
| Cancer | 1388 (18.56) | 497 (17.19) | 307 (13.03) | 396 (30.00) | 188 (20.61) | < 0.001 | - |
| Hyperlipidemia | 3199 (42.77) | 1078 (37.29) | 947 (40.18) | 725 (54.92) | 449 (49.23) | < 0.001 | - |
| Obesity | 3006 (37.30) | 1218 (38.98) | 1018 (39.94) | 484 (34.25) | 286 (29.42) | < 0.001 | - |
| **Outcomes (60 days), N (%)** | | | | | | | |
| Mortality | 1472 (18.27) | 266 (8.51) | 411 (16.12) | 340 (24.06) | 455 (46.81) | < 0.001 | - |
| Mechanical ventilation (intubation) | 1115 (13.84) | 222 (7.10) | 436 (17.10) | 197 (13.94) | 260 (26.75) | < 0.001 | - |
| ICU admission | 1446 (17.94) | 291 (9.31) | 549 (21.54) | 251 (17.76) | 355 (36.52) | < 0.001 | - |
| **Medications, N (%)** | | | | | | | |
| Antibiotics | 2896 (35.93) | 901 (28.83) | 975 (38.25) | 526 (37.23) | 494 (50.82) | < 0.001 | - |
| Corticosteroids | 1628 (20.20) | 415 (13.28) | 595 (23.34) | 294 (20.81) | 324 (33.33) | < 0.001 | - |
| Enoxaparin | 3266 (40.53) | 1203 (38.50) | 1335 (52.37) | 381 (26.96) | 347 (35.70) | < 0.001 | - |
| Heparin | 1280 (15.88) | 357 (11.42) | 472 (18.52) | 283 (20.03) | 168 (17.28) | < 0.001 | - |
| Vasopressor | 582 (7.22) | 138 (4.42) | 248 (9.73) | 100 (7.08) | 96 (9.88) | < 0.001 | - |

Abbreviations: BUN = blood urea nitrogen; COPD = chronic obstructive pulmonary disease; ESR = Erythrocyte sedimentation rate; ICU = intensive care unit; IL-6 = Interleukin 6; IQR = Interquartile range; LDH = Lactate dehydrogenase.

[a] Comparisons across all 4 subphenotypes were performed using the Kruskal-Wallis test (with Dunn's test for post-hoc pairwise comparisons) or $\chi2$ test.

[b] P-values, adjusting for age and sex, were calculated by analysis of covariance (ANCOVA) was performed based on General Linear Model.

[c] 140 patients in development cohort were excluded in the first sensitivity analysis as they had outlier data.

### eTable 7. Characteristics of the subphenotypes in sensitivity analysis 2 (Gaussian mixture model) in the development cohort.

| Variable | Total | Subphenotype I | Subphenotype II | Subphenotype III | Subphenotype IV | P-value[a] | P-value (age and sex adjusted)[b] |
|---|---|---|---|---|---|---|---|
| No. of patients (%) | 8199 (100) | 2641 (32.21) | 2807 (34.24) | 1395 (17.01) | 1356 (16.54) | - | - |
| **Demographics** | | | | | | | |
| Age, y, Median (IQR) | 63.53 (50.57 - 75.15) | 57.05 (42.98 - 70.85) | 62.17 (50.87 - 72.40) | 71.50 (60.21 - 81.02) | 68.59 (57.21 - 79.89) | < 0.001 | - |
| Sex female, N (%) | 3787 (46.19) | 1614 (61.11) | 981 (34.95) | 618 (44.30) | 574 (42.33) | < 0.001 | - |
| Race, N (%) | | | | | | | |
| White | 2036 (24.83) | 684 (25.90) | 741 (26.40) | 325 (23.30) | 286 (21.09) | < 0.001 | - |
| Black | 2155 (26.28) | 720 (27.26) | 521 (18.56) | 488 (34.98) | 426 (31.42) | | |
| Asian | 409 (4.99) | 96 (3.63) | 189 (6.73) | 60 (4.30) | 64 (4.72) | | |
| Other/unknown | 3599 (43.90) | 1141 (43.20) | 1356 (48.31) | 522 (37.42) | 580 (42.77) | | |
| **Inflammatory markers** | | | | | | | |
| C-reactive protein, mg/L, Median (IQR) | 9.40 (3.70 - 16.80) | 3.07 (0.93 - 6.46) | 13.39 (8.20 - 19.32) | 9.05 (4.00 - 15.70) | 13.70 (5.10 - 22.65) | < 0.001 | < 0.001 |
| ESR, mm/hr, Median (IQR) | 69.00 (42.00 - 97.00) | 48.00 (31.00 - 78.50) | 73.00 (50.50 - 97.00) | 76.00 (50.75 - 104.25) | 78.00 (41.00 - 105.00) | < 0.001 | < 0.001 |
| IL-6, pg/mL, Median (IQR) | 19.00 (10.00 - 42.00) | 12.00 (7.00 - 19.77) | 17.00 (10.00 - 41.00) | 21.00 (9.00 - 53.00) | 31.00 (11.00 - 67.00) | < 0.001 | 0.54 |
| Procalcitonin, ng/mL, Median (IQR) | 0.20 (0.10 - 0.60) | 0.10 (0.10 - 0.11) | 0.20 (0.10 - 0.40) | 0.40 (0.20 - 1.06) | 0.50 (0.20 - 1.80) | < 0.001 | 0.001 |
| Bands, %, Median (IQR) | 2.00 (0.00 - 5.00) | 2.00 (0.00 - 5.00) | 2.00 (1.00 - 5.00) | 2.00 (0.00 - 5.00) | 2.00 (0.00 - 6.00) | 0.86 | 0.06 |
| LDH, U/L, Median (IQR) | 377.00 (280.00 - 525.00) | 275.00 (221.75 - 342.00) | 448.00 (367.00 - 584.50) | 353.50 (272.00 - 471.75) | 514.00 (356.00 - 737.00) | < 0.001 | < 0.001 |
| Lymphocyte count, $\times 10^3$/uL, Median (IQR) | 1.00 (0.70 - 1.43) | 1.20 (0.90 - 1.70) | 0.90 (0.60 - 1.20) | 0.80 (0.50 - 1.10) | 1.10 (0.70 - 1.61) | < 0.001 | < 0.001 |
| Neutrophil count, $\times 10^3$/uL, Median (IQR) | 5.30 (3.70 - 7.90) | 3.90 (2.90 - 5.20) | 6.20 (4.50 - 8.40) | 5.00 (3.40 - 7.00) | 9.50 (6.70 - 13.05) | < 0.001 | < 0.001 |
| White blood cell count, $\times 10^3$/uL, Median (IQR) | 7.20 (5.30 - 9.90) | 5.80 (4.60 - 7.40) | 7.80 (6.00 - 10.10) | 6.60 (4.70 - 8.90) | 12.10 (8.80 - 16.30) | < 0.001 | < 0.001 |
| **Inflammation & Hepatic markers** | | | | | | | < 0.001 |
| Albumin, g/dL, Median (IQR) | 3.70 (3.30 - 4.10) | 4.00 (3.70 - 4.40) | 3.70 (3.30 - 4.00) | 3.50 (3.00 - 3.90) | 3.40 (2.80 - 3.80) | < 0.001 | < 0.001 |
| Ferritin, ng/mL, Median (IQR) | 645.00 (295.90 - 1347.00) | 268.00 (129.00 - 470.50) | 925.00 (530.25 - 1543.50) | 713.70 (306.25 - 1599.42) | 957.70 (376.00 - 1919.00) | < 0.001 | < 0.001 |
| **Hepatic markers** | | | | | | | |
| Alanine aminotransferase, U/L, Median (IQR) | 29.00 (19.00 - 48.00) | 23.00 (16.00 - 32.00) | 44.00 (30.00 - 68.00) | 20.00 (13.00 - 28.00) | 36.00 (20.00 - 71.00) | < 0.001 | < 0.001 |
| Aspartate aminotransferase, U/L, Median (IQR) | 39.00 (26.00 - 63.00) | 28.00 (22.00 - 37.00) | 56.00 (41.00 - 81.00) | 32.00 (22.00 - 48.00) | 56.00 (29.00 - 107.75) | < 0.001 | < 0.001 |
| Bilirubin, mg/dL, Median (IQR) | 0.30 (0.20 - 0.60) | 0.20 (0.20 - 0.40) | 0.40 (0.20 - 0.70) | 0.30 (0.20 - 0.50) | 0.40 (0.20 - 0.80) | < 0.001 | < 0.001 |
| **Cardiovascular markers** | | | | | | | |
| Creatine kinase, U/L, Median (IQR) | 154.00 (78.00 - 359.00) | 109.00 (65.75 - 187.25) | 207.50 (101.25 - 463.00) | 141.00 (71.00 - 334.50) | 226.00 (87.00 - 762.75) | < 0.001 | < 0.001 |
| Lactate, mmol/L, Median (IQR) | 1.90 (1.40 - 2.60) | 1.50 (1.20 - 2.00) | 1.90 (1.50 - 2.60) | 1.70 (1.30 - 2.40) | 3.00 (2.00 - 4.50) | < 0.001 | < 0.001 |
| Troponin I, ng/mL, Median (IQR) | 0.10 (0.06 - 0.30) | 0.10 (0.00 - 0.10) | 0.10 (0.04 - 0.20) | 0.10 (0.10 - 0.27) | 0.20 (0.10 - 0.60) | < 0.001 | 0.16 |
| Troponin T, ng/mL, Median (IQR) | 0.01 (0.01 - 0.03) | 0.01 (0.01 - 0.01) | 0.01 (0.01 - 0.01) | 0.04 (0.01 - 0.10) | 0.03 (0.01 - 0.12) | < 0.001 | < 0.001 |
| **Renal markers** | | | | | | | |
| Bicarbonate, mmol/L, Median (IQR) | 23.00 (21.00 - 26.00) | 24.00 (22.00 - 26.00) | 24.00 (21.00 - 26.00) | 22.00 (19.00 - 25.00) | 21.00 (17.00 - 24.00) | < 0.001 | < 0.001 |
| BUN, mg/dL, Median (IQR) | 17.00 (11.00 - 31.00) | 13.00 (9.75 - 17.00) | 16.00 (11.00 - 21.00) | 39.00 (25.00 - 59.00) | 38.00 (19.00 - 71.00) | < 0.001 | < 0.001 |

| | | | | | | | |
|---|---|---|---|---|---|---|---|
| Creatinine, mg/dL, Median (IQR) | 1.00 (0.80 - 1.50) | 0.87 (0.70 - 1.06) | 0.95 (0.80 - 1.20) | 2.20 (1.30 - 4.70) | 1.57 (0.90 - 2.90) | < 0.001 | < 0.001 |
| Chloride, mmol/L, Median (IQR) | 100.00 (97.00 - 104.00) | 102.00 (99.00 - 104.00) | 99.00 (96.00 - 102.00) | 100.00 (95.00 - 105.00) | 103.00 (96.00 - 110.00) | < 0.001 | < 0.001 |
| Sodium, mmol/L, Median (IQR) | 137.00 (134.00 - 140.00) | 138.00 (136.00 - 141.00) | 135.00 (132.00 - 138.00) | 137.00 (134.00 - 140.00) | 139.00 (134.00 - 146.00) | < 0.001 | < 0.001 |
| **Hematologic markers** | | | | | | | |
| D-dimer, ng/mL, Median (IQR) | 1360.00 (620.00 - 3370.00) | 622.00 (370.00 - 1140.00) | 1210.00 (640.00 - 2420.00) | 1940.00 (960.00 - 3610.00) | 3430.00 (1550.00 - 10300.00) | < 0.001 | < 0.001 |
| Hemoglobin, g/dL, Median (IQR) | 13.10 (11.50 - 14.60) | 13.20 (12.10 - 14.40) | 13.90 (12.70 - 15.20) | 10.80 (9.10 - 12.35) | 12.80 (10.60 - 14.90) | < 0.001 | < 0.001 |
| Platelet count, $\times 10^3$/uL, Median (IQR) | 211.00 (162.00 - 277.00) | 206.00 (165.00 - 258.00) | 212.00 (166.00 - 270.00) | 191.00 (139.00 - 263.00) | 257.00 (175.00 - 365.00) | < 0.001 | < 0.001 |
| Prothrombin time, s, Median (IQR) | 13.30 (12.20 - 14.60) | 12.60 (11.90 - 13.60) | 13.40 (12.53 - 14.50) | 13.20 (11.90 - 14.60) | 14.30 (12.70 - 17.20) | < 0.001 | < 0.001 |
| Red blood cell distribution width, %, Median (IQR) | 13.80 (12.90 - 15.00) | 13.50 (12.80 - 14.50) | 13.20 (12.60 - 14.10) | 15.20 (13.90 - 16.90) | 14.90 (13.60 - 17.00) | < 0.001 | < 0.001 |
| Glucose, mg/dL, Median (IQR) | 121.00 (101.00 - 165.00) | 107.00 (95.00 - 127.00) | 127.00 (108.00 - 168.00) | 127.00 (101.00 - 190.00) | 153.00 (112.25 - 258.00) | < 0.001 | < 0.001 |
| **Other markers** | | | | | | | |
| Oxygen saturation, %, Median (IQR) | 69.00 (50.00 - 85.00) | 64.50 (47.50 - 84.90) | 69.00 (51.00 - 85.00) | 70.00 (47.00 - 80.50) | 74.00 (56.00 - 92.00) | 0.03 | 0.06 |
| BMI, kg/m$^2$, Median (IQR) | 28.00 (25.00 - 33.00) | 29.00 (25.00 - 34.00) | 29.00 (25.00 - 33.00) | 27.00 (24.00 - 32.00) | 26.00 (23.00 - 31.00) | < 0.001 | 0.51 |
| **Comorbidity, (missing=590), N (%)** | | | | | | | |
| Hypertension | 4744 (62.35) | 1255 (51.16) | 1480 (56.88) | 1130 (86.00) | 879 (70.89) | < 0.001 | - |
| Diabetes | 3104 (40.79) | 701 (28.58) | 978 (37.59) | 802 (61.04) | 623 (50.24) | < 0.001 | - |
| Coronary artery disease | 1753 (23.04) | 379 (15.45) | 446 (17.14) | 571 (43.46) | 357 (28.79) | < 0.001 | - |
| Heart failure | 1132 (14.88) | 205 (8.36) | 188 (7.23) | 483 (36.76) | 256 (20.65) | < 0.001 | - |
| COPD | 972 (12.77) | 259 (10.56) | 206 (7.92) | 305 (23.21) | 202 (16.29) | < 0.001 | - |
| Asthma | 1091 (14.34) | 415 (16.92) | 301 (11.57) | 223 (16.97) | 152 (12.26) | < 0.001 | - |
| Cancer | 1438 (18.90) | 380 (15.49) | 362 (13.91) | 407 (30.97) | 289 (23.31) | < 0.001 | - |
| Hyperlipidemia | 3262 (42.87) | 851 (34.69) | 1018 (39.12) | 822 (62.56) | 571 (46.05) | < 0.001 | - |
| Obesity | 3039 (37.07) | 1076 (40.74) | 1141 (40.65) | 461 (33.04) | 361 (26.62) | < 0.001 | - |
| **Outcomes (60 days), N (%)** | | | | | | | |
| Mortality | 1529 (18.65) | 141 (5.34) | 482 (17.17) | 382 (27.38) | 524 (38.64) | < 0.001 | - |
| Mechanical ventilation (intubation | 1154 (14.07) | 129 (4.88) | 495 (17.63) | 212 (15.20) | 318 (23.45) | < 0.001 | - |
| ICU admission | 1494 (18.22) | 182 (6.89) | 588 (20.95) | 266 (19.07) | 458 (33.78) | < 0.001 | - |
| **Medications, N (%)** | | | | | | | |
| Antibiotics | 2952 (36.00) | 667 (25.26) | 1117 (39.79) | 577 (41.36) | 591 (43.58) | < 0.001 | - |
| Corticosteroids | 1666 (20.32) | 285 (10.79) | 661 (23.55) | 332 (23.80) | 388 (28.61) | < 0.001 | - |
| Enoxaparin | 3312 (40.40) | 968 (36.65) | 1531 (54.54) | 366 (26.24) | 447 (32.96) | < 0.001 | - |
| Heparin | 1310 (15.98) | 223 (8.44) | 550 (19.59) | 316 (22.65) | 221 (16.30) | < 0.001 | - |
| Vasopressor | 608 (7.42) | 90 (3.41) | 291 (10.37) | 92 (6.59) | 135 (9.96) | < 0.001 | - |

Abbreviations: BUN = blood urea nitrogen; COPD = chronic obstructive pulmonary disease; ESR = Erythrocyte sedimentation rate; ICU = intensive care unit; IL-6 = Interleukin 6; IQR = Interquartile range; LDH = Lactate dehydrogenase.

[a] Comparisons across all 4 subphenotypes were performed using the Kruskal-Wallis test (with Dunn's test for post-hoc pairwise comparisons) or $\chi 2$ test.

[b] P-values, adjusting for age and sex, were calculated by analysis of covariance (ANCOVA) was performed based on General Linear Model.

### eTable 8. Characteristics of the subphenotypes in the internal validation cohort.

| Variable | Total | Subphenotype I | Subphenotype II | Subphenotype III | Subphenotype IV | P-value[a] | P-value (age and sex adjusted)[b] |
|---|---|---|---|---|---|---|---|
| No. of patients (%) | 3519 (100) | 800 (22.73) | 1908 (54.22) | 550 (15.63) | 261 (7.42) | - | - |
| **Demographics** | | | | | | | |
| Age, y, Median (IQR) | 63.51 (50.95 - 75.17) | 58.53 (40.93 - 71.27) | 62.12 (50.72 - 72.42) | 70.82 (59.71 - 80.43) | 73.97 (62.03 - 84.13) | < 0.001 | - |
| Sex female, N (%) | 1585 (45.04) | 478 (59.75) | 737 (38.63) | 262 (47.64) | 108 (41.38) | < 0.001 | - |
| Race, N (%) | | | | | | | |
| White | 838 (23.81) | 206 (25.75) | 464 (24.32) | 122 (22.18) | 46 (17.62) | < 0.001 | - |
| Black | 915 (26.00) | 219 (27.38) | 436 (22.85) | 173 (31.45) | 87 (33.33) | | |
| Asian | 193 (5.48) | 40 (5.00) | 111 (5.82) | 26 (4.73) | 16 (6.13) | | |
| Other/unknown | 1573 (44.70) | 335 (41.87) | 897 (47.01) | 229 (41.63) | 112 (42.91) | | |
| **Inflammatory markers** | | | | | | | |
| C-reactive protein, mg/L, Median (IQR) | 9.63 (4.39 - 16.42) | 1.70 (0.50 - 5.13) | 12.40 (7.43 - 18.50) | 9.61 (4.198 - 16.43) | 13.46 (6.45 - 20.55) | < 0.001 | < 0.001 |
| ESR, mm/hr, Median (IQR) | 64.00 (41.00 - 93.00) | 35.00 (18.50 - 59.00) | 72.00 (53.00 - 95.00) | 66.00 (46.00 - 103.00) | 70.00 (47.50 - 100.25) | < 0.001 | < 0.001 |
| IL-6, pg/mL, Median (IQR) | 16.50 (9.00 - 35.25) | 10.00 (7.00 - 12.00) | 17.00 (9.00 - 32.00) | 16.50 (6.75 - 37.00) | 81.50 (40.00 - 226.50) | < 0.001 | < 0.001 |
| Procalcitonin, ng/mL, Median (IQR) | 0.20 (0.10 - 0.50) | 0.10 (0.10 - 0.20) | 0.20 (0.10 - 0.40) | 0.30 (0.14 - 0.88) | 0.81 (0.30 - 5.67) | < 0.001 | < 0.001 |
| Bands, %, Median (IQR) | 2.00 (1.00 - 6.00) | 3.00 (1.00 - 7.00) | 2.00 (1.00 - 5.00) | 2.00 (0.00 - 4.00) | 3.00 (1.00 - 8.00) | 0.08 | 0.15 |
| LDH, U/L, Median (IQR) | 382.00 (285.00 - 527.00) | 241.00 (202.00 - 295.50) | 410.00 (324.00 - 532.00) | 376.00 (292.00 - 516.00) | 630.00 (474.50 - 885.50) | < 0.001 | < 0.001 |
| Lymphocyte count, $\times 10^3$/uL, Median (IQR) | 1.00 (0.70 - 1.40) | 1.30 (0.90 - 1.90) | 1.00 (0.70 - 1.30) | 0.80 (0.50 - 1.10) | 1.10 (0.80 - 1.70) | < 0.001 | < 0.001 |
| Neutrophil count, $\times 10^3$/uL, Median (IQR) | 5.30 (3.70 - 7.70) | 4.00 (2.70 - 5.70) | 5.60 (4.00 - 7.90) | 5.10 (3.70 - 7.20) | 9.90 (7.20 - 12.93) | < 0.001 | < 0.001 |
| White blood cell count, $\times 10^3$/uL, Median (IQR) | 7.10 (5.30 - 9.60) | 6.10 (4.50 - 8.10) | 7.30 (5.50 - 9.78) | 6.80 (5.00 - 9.10) | 12.10 (8.95 - 15.95) | < 0.001 | < 0.001 |
| **Inflammation & Hepatic markers** | | | | | | | |
| Albumin, g/dL, Median (IQR) | 3.70 (3.30 - 4.10) | 4.10 (3.70 - 4.40) | 3.70 (3.30 - 4.10) | 3.50 (3.00 - 3.80) | 3.40 (2.90 - 3.80) | < 0.001 | < 0.001 |
| Ferritin, ng/mL, Median (IQR) | 699.50 (297.00 - 1435.00) | 223.00 (88.53 - 475.60) | 854.15 (443.25 - 1544.72) | 710.65 (261.00 - 1764.00) | 1275.30 (617.00 - 2535.00) | < 0.001 | < 0.001 |
| **Hepatic markers** | | | | | | | |
| Alanine aminotransferase, U/L, Median (IQR) | 30.00 (19.00 - 49.00) | 20.00 (14.00 - 28.00) | 38.00 (25.00 - 62.00) | 24.00 (15.00 - 34.00) | 40.00 (25.00 - 71.00) | < 0.001 | < 0.001 |
| Aspartate aminotransferase, U/L, Median (IQR) | 40.00 (27.00 - 64.00) | 25.00 (20.00 - 33.00) | 49.00 (34.00 - 75.00) | 37.00 (25.00 - 57.50) | 64.50 (38.25 - 121.00) | < 0.001 | < 0.001 |
| Bilirubin, mg/dL, Median (IQR) | 0.30 (0.20 - 0.60) | 0.30 (0.20 - 0.50) | 0.40 (0.20 - 0.60) | 0.30 (0.20 - 0.50) | 0.30 (0.20 - 0.60) | < 0.001 | < 0.001 |
| **Cardiovascular markers** | | | | | | | |
| Creatine kinase, U/L, Median (IQR) | 151.00 (80.50 - 335.50) | 100.50 (61.00 - 164.00) | 171.00 (89.00 - 361.75) | 135.00 (69.25 - 322.25) | 405.00 (190.00 - 1174.00) | < 0.001 | < 0.001 |
| Lactate, mmol/L, Median (IQR) | 1.90 (1.30 - 2.60) | 1.45 (1.10 - 2.10) | 1.90 (1.40 - 2.60) | 1.80 (1.30 - 2.50) | 3.40 (2.20 - 5.60) | < 0.001 | < 0.001 |
| Troponin I, ng/mL, Median (IQR) | 0.10 (0.07 - 0.30) | 0.07 (0.00 - 0.10) | 0.10 (0.05 - 0.20) | 0.10 (0.10 - 0.30) | 0.27 (0.10 - 0.90) | < 0.001 | 0.09 |
| Troponin T, ng/mL, Median (IQR) | 0.01 (0.01 - 0.02) | 0.01 (0.01 - 0.01) | 0.01 (0.01 - 0.01) | 0.04 (0.01 - 0.14) | 0.07 (0.02 - 0.16) | < 0.001 | < 0.001 |
| **Renal markers** | | | | | | | |
| Bicarbonate, mmol/L, Median (IQR) | 23.00 (21.00 - 26.00) | 24.00 (22.00 - 26.00) | 24.00 (21.00 - 26.00) | 21.00 (19.00 - 25.00) | 19.00 (16.00 - 23.00) | < 0.001 | < 0.001 |
| BUN, mg/dL, Median (IQR) | 17.00 (11.00 - 29.00) | 13.00 (10.00 - 19.00) | 15.00 (11.00 - 22.00) | 40.00 (23.00 - 62.50) | 62.00 (36.00 - 95.00) | < 0.001 | < 0.001 |

| | | | | | | | |
|---|---|---|---|---|---|---|---|
| Creatinine, mg/dL, Median (IQR) | 1.00 (0.80 - 1.50) | 0.85 (0.70 - 1.10) | 0.93 (0.80 - 1.20) | 2.20 (1.20 - 4.80) | 2.42 (1.51 - 4.00) | < 0.001 | < 0.001 |
| Chloride, mmol/L, Median (IQR) | 100.00 (97.00 - 104.00) | 103.00 (100.00 - 105.00) | 99.00 (96.00 - 102.00) | 101.00 (96.00 - 105.00) | 110.00 (100.00 - 120.00) | < 0.001 | < 0.001 |
| Sodium, mmol/L, Median (IQR) | 137.00 (134.00 - 140.00) | 139.00 (137.00 - 141.00) | 136.00 (133.00 - 139.00) | 137.00 (134.00 - 141.00) | 148.00 (139.00 - 159.00) | < 0.001 | < 0.001 |
| **Hematologic markers** | | | | | | | |
| D-dimer, ng/mL, Median (IQR) | 1330.00 (585.25 - 3092.50) | 636.00 (370.00 - 1450.00) | 1210.00 (586.00 - 2740.00) | 1975.00 (1008.00 - 3822.50) | 4405.00 (2025.00 - 13922.50) | < 0.001 | < 0.001 |
| Hemoglobin, g/dL, Median (IQR) | 13.30 (11.70 - 14.70) | 13.10 (11.80 - 14.30) | 13.90 (12.50 - 15.10) | 10.40 (8.50 - 12.30) | 13.50 (11.80 - 15.60) | < 0.001 | < 0.001 |
| Platelet count, $\times 10^3$/uL, Median (IQR) | 210.00 (163.00 - 274.00) | 212.00 (163.75 - 266.00) | 209.00 (166.00 - 271.00) | 205.00 (144.00 - 289.00) | 221.50 (161.25 - 310.00) | 0.14 | 0.002 |
| Prothrombin time, s, Median (IQR) | 13.20 (12.20 - 14.40) | 12.40 (11.60 - 13.60) | 13.40 (12.50 - 14.40) | 13.15 (12.00 - 15.88) | 14.10 (13.10 - 16.40) | < 0.001 | < 0.001 |
| Red blood cell distribution width, %, Median (IQR) | 13.70 (13.00 - 14.90) | 13.70 (13.00 - 14.70) | 13.30 (12.70 - 14.20) | 15.50 (14.10 - 17.80) | 14.70 (13.80 - 16.17) | < 0.001 | < 0.001 |
| Glucose, mg/dL, Median (IQR) | 122.00 (101.50 - 164.00) | 103.00 (91.75 - 120.00) | 125.00 (107.00 - 169.00) | 133.00 (100.00 - 192.00) | 162.00 (126.00 - 258.00) | < 0.001 | < 0.001 |
| **Other markers** | | | | | | | |
| Oxygen saturation, %, Median (IQR) | 72.00 (48.50 - 90.00) | 68.00 (47.00 - 82.75) | 72.00 (48.00 - 86.00) | 76.00 (49.50 - 93.00) | 91.00 (67.00 - 95.50) | 0.06 | 0.11 |
| BMI, kg/m$^2$, Median (IQR) | 28.00 (24.00 - 33.00) | 28.00 (24.00 - 33.00) | 29.00 (25.00 - 33.00) | 27.00 (23.00 - 31.25) | 26.00 (22.00 - 30.00) | < 0.001 | < 0.001 |
| **Comorbidity, (missing=590), N (%)** | | | | | | | |
| Hypertension | 2051 (62.86) | 371 (51.60) | 1053 (58.99) | 428 (84.75) | 199 (78.35) | < 0.001 | - |
| Diabetes | 1288 (39.47) | 187 (26.01) | 674 (37.76) | 302 (59.80) | 125 (49.21) | < 0.001 | - |
| Coronary artery disease | 774 (23.72) | 134 (18.64) | 320 (17.93) | 234 (46.34) | 86 (33.86) | < 0.001 | - |
| Heart failure | 488 (14.96) | 76 (10.57) | 185 (10.36) | 174 (34.46) | 53 (20.87) | < 0.001 | - |
| COPD | 385 (11.80) | 83 (11.54) | 153 (8.57) | 108 (21.39) | 41 (16.14) | < 0.001 | - |
| Asthma | 463 (14.19) | 138 (19.19) | 208 (11.65) | 88 (17.43) | 29 (11.42) | < 0.001 | - |
| Cancer | 575 (17.62) | 125 (17.39) | 243 (13.61) | 156 (30.89) | 51 (20.08) | < 0.001 | - |
| Hyperlipidemia | 1364 (41.80) | 236 (32.82) | 689 (38.60) | 303 (60.00) | 136 (53.54) | < 0.001 | - |
| Obesity | 1261 (35.83) | 282 (35.25) | 743 (38.94) | 171 (31.09) | 65 (24.90) | < 0.001 | - |
| **Outcomes (60 days), N (%)** | | | | | | | |
| Mortality | 696 (19.78) | 37 (4.62) | 343 (17.98) | 173 (31.45) | 143 (54.79) | < 0.001 | - |
| Mechanical ventilation (intubation | 497 (14.12) | 22 (2.75) | 289 (15.15) | 110 (20.00) | 76 (29.12) | < 0.001 | - |
| ICU admission | 661 (18.78) | 49 (6.12) | 382 (20.02) | 130 (23.64) | 100 (38.31) | < 0.001 | - |
| **Medications, N (%)** | | | | | | | |
| Antibiotics | 1234 (35.07) | 183 (22.88) | 734 (38.47) | 186 (33.82) | 131 (50.19) | < 0.001 | - |
| Corticosteroids | 733 (20.83) | 80 (10.00) | 431 (22.59) | 127 (23.09) | 95 (36.40) | < 0.001 | - |
| Enoxaparin | 1403 (39.87) | 242 (30.25) | 947 (49.63) | 141 (25.64) | 73 (27.97) | < 0.001 | - |
| Heparin | 552 (15.69) | 73 (9.12) | 316 (16.56) | 118 (21.45) | 45 (17.24) | < 0.001 | - |
| Vasopressor | 276 (7.84) | 22 (2.75) | 166 (8.70) | 54 (9.82) | 34 (13.03) | < 0.001 | - |

Abbreviations: BUN = blood urea nitrogen; COPD = chronic obstructive pulmonary disease; ESR = Erythrocyte sedimentation rate; ICU = intensive care unit; IL-6 = Interleukin 6; IQR = Interquartile range; LDH = Lactate dehydrogenase.

[a] Comparisons across all 4 subphenotypes were performed using the Kruskal-Wallis test (with Dunn's test for post-hoc pairwise comparisons) or $\chi 2$ test.

[b] P-values, adjusting for age and sex, were calculated by analysis of covariance (ANCOVA) was performed based on General Linear Model.

### eTable 9. Characteristics of the predicted subphenotypes in the external validation cohort.

| Variable | Total | Subphenotype I | Subphenotype II | Subphenotype III | Subphenotype IV | P-value[a] | P-value (age and sex adjusted)[b] |
|---|---|---|---|---|---|---|---|
| No. of patients (%) | 2700 (100) | 918 (34.00) | 929 (34.41) | 499 (18.48) | 354 (13.11) | - | - |
| **Demographics** | | | | | | | |
| Age, y, Median (IQR) | 65.58 (51.08 - 77.39) | 58.52 (39.53 - 71.23) | 65.03 (53.78 - 74.69) | 69.44 (57.23 - 80.55) | 76.53 (68.07 - 85.33) | < 0.001 | - |
| Sex female, N (%) | 1305 (48.33) | 544 (59.26) | 346 (37.24) | 267 (53.51) | 148 (41.81) | < 0.001 | - |
| Race, N (%) | | | | | | | |
| White | 675 (25.00) | 239 (26.03) | 238 (25.62) | 124 (24.85) | 74 (20.90) | < 0.001 | - |
| Black | 545 (20.19) | 186 (20.26) | 136 (14.64) | 135 (27.05) | 88 (24.86) | | |
| Asian | 28 (1.04) | 7 (0.76) | 16 (1.72) | 3 (0.60) | 2 (0.56) | | |
| Other/unknown | 1452 (53.78) | 486 (52.94) | 539 (58.02) | 237 (47.50) | 190 (53.67) | | |
| **Inflammatory markers** | | | | | | | |
| C-reactive protein, mg/L, Median (IQR) | 11.30 (5.50 - 20.15) | 6.16 (1.47 - 11.21) | 15.35 (8.99 - 23.99) | 9.78 (4.46 - 18.33) | 16.29 (8.99 - 27.69) | < 0.001 | < 0.001 |
| ESR, mm/hr, Median (IQR) | 72.00 (50.00 - 97.00) | 56.00 (36.50 - 81.00) | 74.00 (56.00 - 97.00) | 80.00 (55.00 - 103.00) | 86.00 (60.00 - 109.00) | < 0.001 | < 0.001 |
| IL-6, pg/mL, Median (IQR) | 45.20 (11.15 - 108.40) | 10.50 (5.00 - 38.60) | 67.25 (22.95 - 128.93) | 40.30 (18.80 - 97.50) | 134.60 (59.42 - 303.15) | < 0.001 | < 0.001 |
| Procalcitonin, ng/mL, Median (IQR) | 0.20 (0.10 - 0.60) | 0.10 (0.10 - 0.20) | 0.30 (0.10 - 0.50) | 0.30 (0.20 - 1.30) | 0.90 (0.40 - 2.85) | < 0.001 | < 0.001 |
| Bands, %, Median (IQR) | 3.00 (2.00 - 5.00) | 2.00 (1.00 - 4.00) | 3.00 (1.00 - 5.00) | 3.00 (2.00 - 5.25) | 4.00 (2.00 - 6.50) | 0.02 | 0.05 |
| LDH, U/L, Median (IQR) | 419.00 (310.50 - 599.50) | 307.00 (232.00 - 397.50) | 497.00 (381.00 - 689.00) | 365.50 (291.25 - 475.50) | 621.00 (455.00 - 873.00) | < 0.001 | < 0.001 |
| Lymphocyte count, $\times 10^3$/uL, Median (IQR) | 1.00 (0.70 - 1.50) | 1.10 (0.80 - 1.60) | 1.00 (0.80 - 1.40) | 0.80 (0.60 - 1.20) | 1.00 (0.60 - 1.50) | < 0.001 | 0.01 |
| Neutrophil count, $\times 10^3$/uL, Median (IQR) | 5.40 (3.70 - 8.10) | 4.10 (3.00 - 5.40) | 7.00 (4.90 - 9.95) | 4.50 (3.20 - 6.20) | 8.90 (6.50 - 12.75) | < 0.001 | < 0.001 |
| White blood cell count, $\times 10^3$/uL, Median (IQR) | 7.30 (5.40 - 10.20) | 6.20 (4.80 - 7.70) | 8.90 (6.60 - 12.30) | 6.00 (4.60 - 8.00) | 10.95 (8.10 - 15.65) | < 0.001 | < 0.001 |
| **Inflammation & Hepatic markers** | | | | | | | |
| Albumin, g/dL, Median (IQR) | 3.80 (3.40 - 4.10) | 4.00 (3.70 - 4.40) | 3.70 (3.40 - 4.10) | 3.60 (3.20 - 3.92) | 3.40 (3.00 - 3.70) | < 0.001 | < 0.001 |
| Ferritin, ng/mL, Median (IQR) | 672.65 (333.03 - 1276.00) | 346.70 (169.60 - 655.50) | 894.20 (480.60 - 1475.25) | 637.90 (260.00 - 1331.00) | 979.10 (531.45 - 1962.75) | < 0.001 | < 0.001 |
| **Hepatic markers** | | | | | | | |
| Alanine aminotransferase, U/L, Median (IQR) | 27.00 (17.00 - 47.00) | 23.00 (16.00 - 33.00) | 40.00 (23.00 - 67.00) | 18.00 (13.00 - 26.00) | 39.00 (23.00 - 73.00) | < 0.001 | < 0.001 |
| Aspartate aminotransferase, U/L, Median (IQR) | 39.00 (26.00 - 65.00) | 30.00 (22.00 - 42.00) | 55.00 (35.00 - 87.00) | 30.00 (21.00 - 42.00) | 72.00 (43.00 - 134.00) | < 0.001 | < 0.001 |
| Bilirubin, mg/dL, Median (IQR) | 0.30 (0.20 - 0.50) | 0.20 (0.10 - 0.40) | 0.40 (0.20 - 0.60) | 0.20 (0.10 - 0.40) | 0.40 (0.20 - 0.70) | < 0.001 | < 0.001 |
| **Cardiovascular markers** | | | | | | | |
| Creatine kinase, U/L, Median (IQR) | 154.50 (79.00 - 347.75) | 103.00 (61.00 - 208.00) | 166.00 (92.00 - 393.75) | 131.00 (66.50 - 243.00) | 364.00 (135.00 - 1023.00) | < 0.001 | < 0.001 |
| Lactate, mmol/L, Median (IQR) | 1.60 (1.20 - 2.30) | 1.20 (1.00 - 1.60) | 1.70 (1.30 - 2.40) | 1.30 (1.00 - 1.80) | 2.90 (2.00 - 4.60) | < 0.001 | < 0.001 |
| Troponin I, ng/mL, Median (IQR) | 0.00 (0.00 - 0.05) | 0.00 (0.00 - 0.00) | 0.00 (0.00 - 0.00) | 0.00 (0.00 - 0.00) | 0.10 (0.10 - 0.20) | < 0.001 | 0.29 |
| Troponin T, ng/mL, Median (IQR) | 0.02 (0.01 - 0.04) | 0.01 (0.01 - 0.01) | 0.01 (0.01 - 0.03) | 0.04 (0.02 - 0.10) | 0.08 (0.04 - 0.15) | < 0.001 | < 0.001 |
| **Renal markers** | | | | | | | |
| Bicarbonate, mmol/L, Median (IQR) | 23.00 (20.00 - 25.00) | 24.00 (22.00 - 26.00) | 23.00 (20.00 - 25.00) | 22.00 (19.00 - 25.00) | 19.00 (16.00 - 22.00) | < 0.001 | < 0.001 |
| BUN, mg/dL, Median (IQR) | 18.00 (12.00 - 33.00) | 12.00 (9.00 - 16.00) | 17.00 (12.00 - 26.00) | 35.00 (22.00 - 57.00) | 54.00 (34.00 - 83.00) | < 0.001 | < 0.001 |

| | | | | | | | |
|---|---|---|---|---|---|---|---|
| Creatinine, mg/dL, Median (IQR) | 1.10 (0.80 - 1.60) | 0.90 (0.70 - 1.10) | 1.00 (0.80 - 1.30) | 2.10 (1.20 - 4.90) | 2.10 (1.30 - 3.50) | < 0.001 | < 0.001 |
| Chloride, mmol/L, Median (IQR) | 100.00 (96.00 - 103.00) | 100.00 (98.00 - 103.00) | 98.00 (95.00 - 101.00) | 101.00 (96.00 - 104.00) | 105.00 (98.00 - 114.00) | < 0.001 | < 0.001 |
| Sodium, mmol/L, Median (IQR) | 138.00 (135.00 - 141.00) | 138.00 (136.00 - 141.00) | 137.00 (134.00 - 139.00) | 138.00 (135.00 - 141.00) | 143.00 (137.00 - 153.50) | < 0.001 | < 0.001 |
| **Hematologic markers** | | | | | | | |
| D-dimer, ng/mL, Median (IQR) | 1400.00 (700.00 - 3100.00) | 750.00 (400.00 - 1400.00) | 1500.00 (900.00 - 2800.00) | 1900.00 (1100.00 - 3400.00) | 4250.00 (2325.00 - 13875.00) | < 0.001 | < 0.001 |
| Hemoglobin, g/dL, Median (IQR) | 13.00 (11.40 - 14.30) | 13.20 (12.10 - 14.30) | 13.70 (12.50 - 14.80) | 10.60 (9.30 - 11.90) | 12.70 (10.80 - 14.45) | < 0.001 | < 0.001 |
| Platelet count, $\times 10^3$/uL, Median (IQR) | 203.00 (157.00 - 262.50) | 198.00 (158.00 - 248.00) | 216.00 (170.00 - 290.00) | 179.00 (139.00 - 240.00) | 210.00 (159.00 - 289.00) | < 0.001 | < 0.001 |
| Prothrombin time, s, Median (IQR) | 14.20 (13.40 - 15.30) | 13.60 (13.10 - 14.30) | 14.10 (13.40 - 15.00) | 14.45 (13.60 - 15.57) | 16.10 (14.80 - 19.20) | < 0.001 | < 0.001 |
| Red blood cell distribution width, %, Median (IQR) | 13.90 (13.00 - 15.00) | 13.50 (12.80 - 14.30) | 13.40 (12.80 - 14.30) | 15.30 (14.00 - 17.20) | 15.00 (13.90 - 16.40) | < 0.001 | < 0.001 |
| Glucose, mg/dL, Median (IQR) | 125.00 (104.00 - 176.00) | 108.00 (96.00 - 126.00) | 138.00 (115.00 - 219.75) | 122.00 (100.25 - 156.00) | 183.00 (127.00 - 288.00) | < 0.001 | < 0.001 |
| **Other markers** | | | | | | | |
| Oxygen saturation, %, Median (IQR) | 97.10 (92.00 - 98.70) | 95.25 (73.88 - 98.45) | 96.80 (93.00 - 98.70) | 97.00 (92.40 - 98.80) | 98.00 (91.60 - 98.82) | 0.70 | 0.23 |
| BMI, kg/m$^2$, Median (IQR) | 28.00 (24.00 - 33.00) | 29.00 (25.00 - 34.00) | 28.00 (25.00 - 32.00) | 26.50 (23.00 - 32.00) | 26.00 (22.00 - 30.00) | < 0.001 | 0.65 |
| **Comorbidity, (missing=590), N (%)** | | | | | | | |
| Hypertension | 539 (20.01) | 131 (14.35) | 181 (19.48) | 151 (30.32) | 76 (21.47) | < 0.001 | - |
| Diabetes | 438 (16.26) | 82 (8.98) | 169 (18.19) | 121 (24.30) | 66 (18.64) | < 0.001 | - |
| Coronary artery disease | 167 (6.20) | 26 (2.85) | 50 (5.38) | 64 (12.85) | 27 (7.63) | < 0.001 | - |
| Heart failure | 153 (5.68) | 18 (1.97) | 39 (4.20) | 60 (12.05) | 36 (10.17) | < 0.001 | - |
| COPD | 74 (2.75) | 17 (1.86) | 28 (3.01) | 18 (3.61) | 11 (3.11) | 0.21 | - |
| Asthma | 101 (3.75) | 51 (5.59) | 31 (3.34) | 14 (2.81) | 5 (1.41) | 0.001 | - |
| Cancer | 175 (6.50) | 51 (5.59) | 55 (5.92) | 45 (9.04) | 24 (6.78) | 0.07 | - |
| Hyperlipidemia | 193 (7.16) | 42 (4.60) | 72 (7.75) | 49 (9.84) | 30 (8.47) | 0.001 | - |
| Obesity | 910 (33.70) | 339 (36.93) | 327 (35.20) | 160 (32.06) | 84 (23.73) | < 0.001 | - |
| **Outcomes (60 days), N (%)** | | | | | | | |
| Mortality | 556 (20.59) | 56 (6.10) | 204 (21.96) | 112 (22.44) | 184 (51.98) | < 0.001 | - |
| Mechanical ventilation (intubation | 248 (9.19) | 38 (4.14) | 113 (12.16) | 40 (8.02) | 57 (16.10) | < 0.001 | - |
| ICU admission | - | - | - | - | - | - | - |
| **Medications, N (%)** | | | | | | | |
| Antibiotics | 1739 (64.41) | 450 (49.02) | 655 (70.51) | 351 (70.34) | 283 (79.94) | < 0.001 | - |
| Corticosteroids | 741 (27.44) | 176 (19.17) | 318 (34.23) | 161 (32.26) | 86 (24.29) | < 0.001 | - |
| Enoxaparin | 1788 (66.22) | 604 (65.80) | 744 (80.09) | 271 (54.31) | 169 (47.74) | < 0.001 | - |
| Heparin | 978 (36.22) | 188 (20.48) | 305 (32.83) | 291 (58.32) | 194 (54.80) | < 0.001 | - |
| Vasopressor | 219 (8.11) | 23 (2.51) | 103 (11.09) | 41 (8.22) | 52 (14.69) | < 0.001 | - |

Abbreviations: BUN = blood urea nitrogen; COPD = chronic obstructive pulmonary disease; ESR = Erythrocyte sedimentation rate; ICU = intensive care unit; IL-6 = Interleukin 6; IQR = Interquartile range; LDH = Lactate dehydrogenase.

[a] Comparisons across all 4 subphenotypes were performed using the Kruskal-Wallis test (with Dunn's test for post-hoc pairwise comparisons) or $\chi 2$ test.

[b] P-values, adjusting for age and sex, were calculated by analysis of covariance (ANCOVA) was performed based on General Linear Model.

### eTable 10. Characteristics of the subphenotypes identified in the leave-one-center-out procedure among all studied patients.

| Variable | Total | Subphenotype I | Subphenotype II | Subphenotype III | Subphenotype IV | P-value[a] | P-value (age and sex adjusted)[b] |
|---|---|---|---|---|---|---|---|
| No. of patients (%) | 14418 (100) | 5319 (36.89) | 4627 (32.09) | 3454 (23.96) | 1018 (7.06) | - | - |
| **Demographics** | | | | | | | |
| Age, y, Median (IQR) | 64.01 (50.72 - 75.55) | 59.36 (43.89 - 71.73) | 62.41 (51.48 - 72.59) | 71.66 (58.82 - 81.78) | 68.57 (58.91 - 77.87) | < 0.001 | - |
| Sex female, N (%) | 6677 (46.31) | 2849 (53.56) | 1741 (37.63) | 1719 (49.77) | 368 (36.15) | < 0.001 | - |
| Race, N (%) | | | | | | | |
| White | 3549 (24.62) | 1384 (26.02) | 1117 (24.14) | 833 (24.12) | 215 (21.12) | < 0.001 | - |
| Black | 3615 (25.07) | 1324 (24.89) | 885 (19.13) | 1104 (31.96) | 302 (29.67) | | |
| Asian | 630 (4.37) | 191 (3.59) | 279 (6.03) | 119 (3.45) | 41 (4.03) | | |
| Other/unknown | 6624 (45.95) | 2420 (52.94) | 2346 (50.7) | 1398 (40.48) | 460 (45.19) | | |
| **Inflammatory markers** | | | | | | | |
| C-reactive protein, mg/L, Median (IQR) | 9.90 (4.10 - 174.00) | 4.93 (1.32 - 10.50) | 13.42 (7.60 - 20.44) | 10.30 (4.60 - 17.70) | 16.80 (9.39 - 26.24) | < 0.001 | < 0.001 |
| ESR, mm/hr, Median (IQR) | 69.00 (45.00 - 96.25) | 55.00 (34.00 - 80.00) | 75.00 (54.00 - 99.00) | 76.00 (48.75 - 104.00) | 82.00 (53.00 - 106.00) | < 0.001 | < 0.001 |
| IL-6, pg/mL, Median (IQR) | 22.00 (10.00 - 57.83) | 13.00 (6.00 - 29.00) | 21.00 (11.00 - 55.00) | 33.00 (13.00 - 80.10) | 60.50 (20.75 - 156.18) | < 0.001 | 0.002 |
| Procalcitonin, ng/mL, Median (IQR) | 0.20 (0.10 - 0.60) | 0.10 (0.10 - 0.20) | 0.20 (0.10 - 0.46) | 0.40 (0.20 - 1.30) | 0.70 (0.30 - 2.30) | < 0.001 | < 0.001 |
| Bands, %, Median (IQR) | 2.00 (1.00 - 5.00) | 3.00 (1.00 - 6.00) | 2.00 (0.00 - 5.00) | 2.30 (0.30 - 6.00) | 3.00 (1.00 - 6.00) | 0.02 | 0.01 |
| LDH, U/L, Median (IQR) | 390.00 (288.00 - 542.00) | 299.00 (232.00 - 391.50) | 443.00 (350.00 - 585.25) | 402.00 (301.00 - 562.50) | 645.00 (429.50 - 908.50) | < 0.001 | < 0.001 |
| Lymphocyte count, $\times10^3$/uL, Median (IQR) | 1.00 (0.70 - 1.40) | 1.10 (0.80 - 1.60) | 1.00 (0.70 - 1.30) | 1.00 (0.60 - 1.40) | 0.90 (0.60 - 1.30) | < 0.001 | 0.01 |
| Neutrophil count, $\times10^3$/uL, Median (IQR) | 5.40 (3.70 - 7.90) | 4.00 (2.90 - 5.40) | 6.40 (4.70 - 8.90) | 6.00 (4.10 - 8.80) | 9.20 (6.20 - 13.10) | < 0.001 | < 0.001 |
| White blood cell count, $\times10^3$/uL, Median (IQR) | 7.20 (5.30 - 9.90) | 5.80 (4.50 - 7.50) | 8.20 (6.30 - 10.70) | 7.80 (5.70 - 11.00) | 11.30 (7.80 - 15.70) | < 0.001 | < 0.001 |
| **Inflammation & Hepatic markers** | | | | | | | |
| Albumin, g/dL, Median (IQR) | 3.70 (3.30 - 4.10) | 4.00 (3.60 - 4.30) | 3.60 (3.20 - 4.00) | 3.50 (3.00 - 3.90) | 3.40 (3.00 - 3.80) | < 0.001 | < 0.001 |
| Ferritin, ng/mL, Median (IQR) | 661.00 (303.00 - 1352.25) | 372.00 (172.00 - 772.02) | 846.75 (454.00 - 1506.50) | 722.25 (289.50 - 1536.00) | 1265.00 (612.92 - 2463.50) | < 0.001 | < 0.001 |
| **Hepatic markers** | | | | | | | |
| Alanine aminotransferase, U/L, Median (IQR) | 29.00 (18.00 - 48.00) | 25.00 (17.00 - 38.00) | 39.00 (25.00 - 64.00) | 23.00 (15.00 - 37.00) | 39.00 (22.00 - 83.00) | < 0.001 | < 0.001 |
| Aspartate aminotransferase, U/L, Median (IQR) | 40.00 (26.00 - 64.00) | 32.00 (23.00 - 47.00) | 50.00 (34.00 - 75.00) | 37.00 (24.00 - 61.00) | 67.50 (36.00 - 146.25) | < 0.001 | < 0.001 |
| Bilirubin, mg/dL, Median (IQR) | 0.30 (0.20 - 0.60) | 0.30 (0.20 - 0.50) | 0.40 (0.20 - 0.60) | 0.30 (0.20 - 0.50) | 0.40 (0.20 - 0.80) | < 0.001 | < 0.001 |
| **Cardiovascular markers** | | | | | | | |
| Creatine kinase, U/L, Median (IQR) | 153.00 (79.00 - 349.75) | 124.00 (72.00 - 235.00) | 159.00 (83.25 - 358.75) | 176.00 (77.00 - 493.25) | 293.00 (115.00 - 890.75) | < 0.001 | < 0.001 |
| Lactate, mmol/L, Median (IQR) | 1.80 (1.30 - 2.60) | 1.50 (1.10 - 2.10) | 1.90 (1.50 - 2.60) | 1.90 (1.30 - 2.80) | 3.20 (2.10 - 5.00) | < 0.001 | < 0.001 |
| Troponin I, ng/mL, Median (IQR) | 0.10 (0.04 - 0.27) | 0.10 (0.00 - 0.10) | 0.10 (0.03 - 0.20) | 0.12 (0.10 - 0.37) | 0.30 (0.10 - 1.10) | < 0.001 | < 0.001 |
| Troponin T, ng/mL, Median (IQR) | 0.01 (0.01 - 0.03) | 0.01 (0.01 - 0.01) | 0.01 (0.01 - 0.01) | 0.05 (0.02 - 0.14) | 0.03 (0.01 - 0.10) | < 0.001 | < 0.001 |
| **Renal markers** | | | | | | | |
| Bicarbonate, mmol/L, Median (IQR) | 23.00 (21.00 - 26.00) | 24.00 (22.00 - 26.00) | 24.00 (21.00 - 26.00) | 22.00 (19.00 - 25.00) | 19.00 (16.00 - 22.00) | < 0.001 | < 0.001 |
| BUN, mg/dL, Median (IQR) | 17.00 (11.00 - 31.00) | 13.00 (10.00 - 18.00) | 15.00 (11.00 - 22.00) | 41.00 (22.00 - 69.00) | 39.00 (25.00 - 62.50) | < 0.001 | < 0.001 |

| | | | | | | | |
|---|---|---|---|---|---|---|---|
| Creatinine, mg/dL, Median (IQR) | 1.00 (0.80 - 1.50) | 0.90 (0.70 - 1.10) | 0.92 (0.80 - 1.20) | 2.06 (1.10 - 4.46) | 1.75 (1.20 - 2.80) | < 0.001 | < 0.001 |
| Chloride, mmol/L, Median (IQR) | 100.00 (97.00 - 104.00) | 101.00 (98.00 - 104.00) | 98.00 (95.00 - 101.00) | 103.00 (98.00 - 109.00) | 99.00 (95.00 - 104.00) | < 0.001 | < 0.001 |
| Sodium, mmol/L, Median (IQR) | 137.00 (134.00 - 140.00) | 138.00 (136.00 - 140.00) | 135.00 (132.00 - 138.00) | 139.00 (136.00 - 144.00) | 137.00 (133.00 - 141.00) | < 0.001 | < 0.001 |
| **Hematologic markers** | | | | | | | |
| D-dimer, ng/mL, Median (IQR) | 1370.00 (630.00 - 3250.00) | 770.00 (410.00 - 1500.00) | 1300.00 (660.00 - 2710.00) | 2530.00 (1110.25 - 5843.00) | 3740.00 (1920.00 - 11840.00) | < 0.001 | < 0.001 |
| Hemoglobin, g/dL, Median (IQR) | 13.10 (11.50 - 14.60) | 13.40 (12.12 - 14.70) | 13.80 (12.60 - 15.00) | 11.20 (9.50 - 13.00) | 13.10 (11.10 - 14.90) | < 0.001 | < 0.001 |
| Platelet count, $\times 10^3$/uL, Median (IQR) | 209.00 (162.00 - 273.00) | 196.00 (156.00 - 247.00) | 226.00 (175.00 - 298.00) | 206.00 (153.00 - 278.00) | 228.00 (169.00 - 308.00) | < 0.001 | < 0.001 |
| Prothrombin time, s, Median (IQR) | 13.60 (12.60 - 14.80) | 13.20 (12.30 - 14.00) | 13.60 (12.70 - 14.70) | 14.20 (12.70 - 16.10) | 14.60 (13.20 - 16.80) | < 0.001 | < 0.001 |
| Red blood cell distribution width, %, Median (IQR) | 13.80 (13.00 - 15.00) | 13.45 (12.80 - 14.40) | 13.30 (12.70 - 14.20) | 15.30 (14.10 - 17.20) | 14.30 (13.30 - 15.90) | < 0.001 | < 0.001 |
| Glucose, mg/dL, Median (IQR) | 122.00 (102.00 - 166.00) | 108.00 (96.00 - 127.00) | 135.00 (111.00 - 202.00) | 126.00 (101.00 - 172.00) | 213.00 (141.00 - 354.00) | < 0.001 | < 0.001 |
| **Other markers** | | | | | | | |
| Oxygen saturation, %, Median (IQR) | 78.00 (54.00 - 93.60) | 70.00 (49.00 - 90.00) | 74.00 (54.00 - 92.00) | 80.00 (55.00 - 95.15) | 91.00 (70.00 - 98.00) | < 0.001 | < 0.001 |
| BMI, kg/m$^2$, Median (IQR) | 28.00 (24.00 - 33.00) | 29.00 (25.00 - 33.00) | 28.00 (25.00 - 33.00) | 27.00 (23.00 - 32.00) | 27.00 (24.00 - 32.00) | < 0.001 | 0.59 |
| **Comorbidity, (missing=590), N (%)** | | | | | | | |
| Hypertension | 7334 (54.06) | 2284 (45.29) | 2230 (52.01) | 2194 (67.32) | 626 (64.14) | < 0.001 | - |
| Diabetes | 4830 (35.60) | 1198 (23.76) | 1664 (38.81) | 1432 (43.94) | 536 (54.92) | < 0.001 | - |
| Coronary artery disease | 2694 (19.86) | 695 (13.78) | 643 (15.00) | 1101 (33.78) | 255 (26.13) | < 0.001 | - |
| Heart failure | 1773 (13.07) | 425 (8.43) | 325 (7.58) | 841 (25.81) | 182 (18.65) | < 0.001 | - |
| COPD | 1431 (10.55) | 464 (9.20) | 311 (7.25) | 536 (16.45) | 120 (12.30) | < 0.001 | - |
| Asthma | 1655 (12.20) | 708 (14.04) | 461 (10.75) | 390 (11.97) | 96 (9.84) | < 0.001 | - |
| Cancer | 2188 (16.13) | 722 (14.32) | 553 (12.90) | 748 (22.95) | 165 (16.91) | < 0.001 | - |
| Hyperlipidemia | 4819 (35.52) | 1451 (28.77) | 1481 (34.54) | 1476 (45.29) | 411 (42.11) | < 0.001 | - |
| Obesity | 5210 (36.14) | 2070 (38.91) | 1718 (37.13) | 1071 (31.01) | 351 (34.48) | < 0.001 | - |
| **Outcomes (60 days), N (%)** | | | | | | | |
| Mortality | 2781 (19.29) | 488 (9.17) | 764 (16.51) | 1036 (29.99) | 493 (48.43) | < 0.001 | - |
| Mechanical ventilation (intubation | 1899 (13.17) | 374 (7.03) | 716 (15.47) | 503 (14.56) | 306 (30.06) | < 0.001 | - |
| ICU admission | 2155 (14.95) | 424 (7.97) | 830 (17.94) | 586 (16.97) | 315 (30.94) | <0.001 | - |
| **Medications, N (%)** | | | | | | | |
| Antibiotics | 5925 (41.09) | 1816 (34.14) | 1966 (42.49) | 1529 (44.27) | 614 (60.31) | < 0.001 | - |
| Corticosteroids | 3140 (21.78) | 866 (16.28) | 1116 (24.12) | 813 (23.54) | 345 (33.89) | < 0.001 | - |
| Enoxaparin | 6503 (45.10) | 2399 (45.10) | 2541 (54.92) | 1154 (33.41) | 409 (40.18) | < 0.001 | - |
| Heparin | 2840 (19.70) | 756 (14.21) | 899 (19.43) | 891 (25.80) | 294 (28.88) | < 0.001 | - |
| Vasopressor | 1103 (7.65) | 256 (4.81) | 431 (9.31) | 273 (7.90) | 143 (14.05) | < 0.001 | - |

Abbreviations: BUN = blood urea nitrogen; COPD = chronic obstructive pulmonary disease; ESR = Erythrocyte sedimentation rate; ICU = intensive care unit; IL-6 = Interleukin 6; IQR = Interquartile range; LDH = Lactate dehydrogenase.

[a] Comparisons across all 4 subphenotypes were performed using the Kruskal-Wallis test (with Dunn's test for post-hoc pairwise comparisons) or χ2 test.

[b] P-values, adjusting for age and sex, were calculated by analysis of covariance (ANCOVA) was performed based on General Linear Model.

### eTable 11. Distributions of SDoH variables by subphenotypes in the development cohort

| SDoH variables | Total | Subphenotype I | Subphenotype II | Subphenotype III | Subphenotype IV | P-value[a] |
|---|---|---|---|---|---|---|
| # of patients | 8199 (100) | 2707 (33.02) | 3047 (37.16) | 1486 (18.12) | 959 (11.70) | - |
| Median household income, $, Mean (IQR) | 524090 (37015 - 71225) | 52052 (37015 - 71437) | 52409 (40733 - 71477) | 52052 (37243 - 69799) | 51477 (37015 - 63652) | 0.006 |
| Crowding housing, %, Mean (IQR) | 9.30 (6.00 - 14.48) | 9.30 (6.00 - 14.10) | 9.80 (6.40 - 15.70) | 9.30 (6.00 - 13.50) | 9.80 (6.40 - 15.90) | 0.01 |
| No high school degree, %, Mean (IQR) | 20.49 (14.16 - 27.53) | 20.49 (14.16 - 27.53) | 20.49 (14.16 - 27.53) | 20.49 (14.16 - 26.11) | 20.49 (15.75 - 26.10) | 0.34 |
| Essential occupation rate, %, Mean (IQR) | 44.12 (37.42 - 47.57) | 43.87 (37.32 - 47.57) | 43.87 (37.32 - 47.45) | 45.28 (37.42 - 47.57) | 46.30 (39.65 - 47.57) | <0.001 |
| Non-white resident rate, %, Mean (IQR) | 72.32 (42.69 - 84.25) | 72.32 (41.90 - 84.64) | 69.67 (41.95 - 82.27) | 76.81 (47.29 - 84.64) | 77.38 (51.80 - 84.25) | <0.001 |
| Unemployed rate, %, Mean (IQR) | 7.80 (5.70 - 10.05) | 7.80 (5.70 - 10.05) | 7.20 (5.50 - 10.10) | 7.80 (5.80 - 10.30) | 8.80 (6.50 - 10.70) | <0.001 |

Abbreviations: IQR = Interquartile range; LDH = Lactate dehydrogenase; SDoH = social determinants of health.

[a] Comparisons across all 4 subphenotypes were performed using the Kruskal-Wallis test (with Dunn's test for post-hoc pairwise comparisons) or $\chi 2$ test.

**eTable 12. Log odds ratio by logistic regression analyses showing associations between SDoH variables and 60-day mortality risk in each subphenotype in the development cohort**

| SDoH variables | Subphenotype I | | Subphenotype II | | Subphenotype III | | Subphenotype IV | |
|---|---|---|---|---|---|---|---|---|
| | Log odds (SD) | P-value | Log odds (SD) | P-value | Log odds (SD) | P-value | Log odds (SD) | P-value |
| *Logistic regression analysis adjusting for age and sex.* | | | | | | | | |
| Median household income | -0.09 (0.08) | 0.28 | -0.06 (0.05) | 0.20 | -0.13 (0.07) | 0.06 | -0.14 (0.08) | 0.07 |
| Crowding housing | 0.05 (0.08) | 0.51 | 0.09 (0.04) | 0.05 | 0.07 (0.07) | 0.30 | 0.11 (0.08) | 0.12 |
| No high school degree | 0.09 (0.08) | 0.27 | 0.11 (0.05) | 0.03 | 0.13 (0.06) | 0.05 | 0.14 (0.07) | 0.06 |
| Essential occupation rate | 0.07 (0.08) | 0.40 | 0.12 (0.05) | 0.02 | 0.18 (0.07) | 0.01 | 0.15 (0.07) | 0.04 |
| Non-white resident rate | 0.02 (0.08) | 0.77 | 0.09 (0.05) | 0.10 | 0.07 (0.06) | 0.29 | 0.18 (0.07) | 0.01 |
| Unemployed rate | -0.04 (0.08) | 0.63 | 0.03 (0.05) | 0.53 | 0.11 (0.07) | 0.10 | 0.19 (0.07) | 0.01 |
| *Logistic regression analysis adjusting for age, sex, and clinical variables* | | | | | | | | |
| Median household income | -0.06 (0.09) | 0.51 | 0.01 (0.06) | 0.76 | -0.13 (0.08) | 0.10 | -0.03 (0.09) | 0.73 |
| Crowding housing | -0.01 (0.09) | 0.87 | 0.03 (0.05) | 0.55 | 0.08 (0.07) | 0.28 | 0.02 (0.08) | 0.77 |
| No high school degree | 0.05 (0.09) | 0.57 | 0.04 (0.05) | 0.43 | 0.13 (0.07) | 0.07 | 0.05 (0.08) | 0.53 |
| Essential occupation rate | 0.08 (0.09) | 0.39 | 0.08 (0.06) | 0.15 | 0.16 (0.08) | 0.04 | 0.08 (0.09) | 0.34 |
| Non-white resident rate | -0.025 (0.09) | 0.79 | 0.07 (0.06) | 0.22 | 0.05 (0.08) | 0.51 | 0.13 (0.09) | 0.13 |
| Unemployed rate | -0.11 (0.10) | 0.26 | -0.04 (0.06) | 0.51 | 0.06 (0.07) | 0.28 | 0.10 (0.08) | 0.25 |

Logistic regression analyses were performed the associations between each SDoH variable with 60-day mortality risk in each subphenotype, adjusting for age and sex or for age, sex, and clinical variables.
Abbreviations: SD = standard deviation.

**eTable 13. Hazard ratio by Cox regression analyses showing associations between SDoH variables and 60-day mortality risk in each subphenotype in the development cohort**

| SDoH variables | Subphenotype I | | Subphenotype II | | Subphenotype III | | Subphenotype IV | |
|---|---|---|---|---|---|---|---|---|
| | HR (95% CI) | P-value | HR (95% CI) | P-value | HR (95% CI) | P-value | HR (95% CI) | P-value |
| Median household income | 1.02 (0.89 - 1.16) | 0.83 | 0.97 (0.89 - 1.06) | 0.53 | 0.92 (0.82 - 1.03) | 0.13 | 0.97 (0.87 - 1.08) | 0.59 |
| Crowding housing | 1.01 (0.87 - 1.17) | 0.89 | 1.09 (1.00 - 1.19) | 0.04 | 1.06 (0.95 - 1.18) | 0.31 | 1.05 (0.95 - 1.15) | 0.33 |
| No high school degree | 1.01 (0.88 - 1.17) | 0.85 | 1.08 (1.00 - 1.18) | 0.06 | 1.09 (0.98 - 1.21) | 0.10 | 1.06 (0.95 - 1.17) | 0.06 |
| Essential occupation rate | 1.02 (0.88 - 1.17) | 0.80 | 1.08 (0.99 - 1.18) | 0.07 | 1.12 (1.00 - 1.25) | 0.04 | 1.07 (0.97 - 1.19) | 0.20 |
| Non-white resident rate | 0.96 (0.84 - 1.11) | 0.59 | 1.06 (0.97 - 1.15) | 0.21 | 1.03 (0.92 - 1.14) | 0.62 | 1.08 (0.98 - 1.20) | 0.11 |
| Unemployed rate | 0.88 (0.75 - 1.02) | 0.09 | 0.98 (0.90 - 1.07) | 0.61 | 1.06 (0.95 - 1.18) | 0.27 | 1.09 (0.99 - 1.20) | 0.07 |

Cox regression analyses were performed the associations between each SDoH variable with 60-day mortality risk in each subphenotype, adjusting for age and sex.
Abbreviations: HR = Hazard ratio; CI = confidence interval.

### eTable 14. Characteristics of the identified SDoH strata in the development cohort

| Variable | Total | Stratum H | Stratum M | Stratum L | P-value[a] | P-value (age and sex adjusted)[b] |
|---|---|---|---|---|---|---|
| No. of patients, N (%) | 7862 (100)[c] | 1933 (24.59) | 3912 (49.76) | 2017 (25.66) | - | |
| **Demographics** | | | | | | |
| Age, Median (IQR) | 63.58 (50.51 - 75.16) | 64.76 (50.70 - 76.33) | 63.99 (51.34 - 75.33) | 62.10 (49.11 - 73.83) | < 0.001 | < 0.001 |
| Sex female, N (%) | 3649 (46.41) | 867 (44.85) | 1848 (47.24) | 934 (46.31) | 0.22 | - |
| **COVID confirmation date** | | | | | | |
| Time of COVID confirmation since March 1st, 2020, Day Median (IQR) | 35 (27 - 47) | 35 (26 - 47) | 35 (28 - 46) | 36 (28 - 49) | 0.001 | 0.005 |
| **Neighborhood conditions** | | | | | | |
| Median household income, $, Mean (IQR) | 52409.00 (37015.00 - 71225.00) | 89466.00 (72314.00 - 117069.00) | 52409.00 (50108.00 - 63652.00) | 36046.00 (28277.00 - 37015.00) | < 0.001 | < 0.001 |
| Crowding housing, %, Mean (IQR) | 9.30 (6.00 - 14.48) | 3.70 (2.10 - 7.70) | 9.30 (6.50 - 10.90) | 15.90 (15.90 - 18.10) | < 0.001 | < 0.001 |
| No high school degree, %, Mean (IQR) | 20.49 (14.16 - 27.53) | 8.90 (5.42 - 13.85) | 20.49 (18.86 - 24.34) | 34.24 (26.10 - 37.57) | < 0.001 | < 0.001 |
| Essential occupation rate, %, Mean (IQR) | 44.12 (37.42 - 47.57) | 32.34 (27.77 - 37.19) | 44.24 (40.70 - 47.57) | 47.45 (46.50 - 52.67) | < 0.001 | < 0.001 |
| Non-white resident rate, %, Mean (IQR) | 72.32 (42.69 - 84.25) | 26.82 (22.03 - 35.93) | 76.81 (58.45 - 84.70) | 82.27 (77.38 - 86.81) | < 0.001 | < 0.001 |
| Unemployed rate, %, Mean (IQR) | 7.80 (5.70 - 10.50) | 4.90 (4.00 - 5.90) | 8.20 (6.80 - 9.40) | 12.10 (10.70 - 12.50) | < 0.001 | < 0.001 |
| **60-day outcomes** | | | | | | |
| Mortality | 1522 (19.36) | 340 (17.59) | 779 (19.91) | 403 (19.98) | 0.08 | - |
| Mechanical ventilation (intubation | 1148 (14.60) | 298 (15.42) | 556 (14.21) | 294 (14.58) | 0.47 | - |
| ICU admission | 1459 (18.56) | 416 (21.52) | 717 (18.33) | 326 (16.16) | < 0.001 | - |

Abbreviations: IQR = Interquartile range; LDH = Lactate dehydrogenase; SDoH = social determinants of health.

[a] Comparisons across all 4 subphenotypes were performed using the Kruskal-Wallis test (with Dunn's test for post-hoc pairwise comparisons) or χ2 test.

[b] P-values, adjusting for age and sex, were calculated by analysis of covariance (ANCOVA) was performed based on General Linear Model.

[c] Of 8199 patients in the development cohort, 7862 patients containing SDoH data were used to derive SDoH strata.

### eTable 15. Distributions of SDoH strata by subphenotypes in the development cohort

| SDoH strata | Total | Subphenotype I | Subphenotype II | Subphenotype III | Subphenotype IV | P-value[a] |
|---|---|---|---|---|---|---|
| # of patients | 7862 | 2593 | 2905 | 1432 | 932 | - |
| Stratum H | 1933 (24.59%) | 690 (26.61%) | 732 (25.20%) | 331 (23.11%) | 180 (19.31%) | < 0.001 |
| Stratum M | 3912 (49.76%) | 1230 (47.44%) | 1441 (49.60%) | 759 (53.00%) | 482 (51.72%) | |
| Stratum L | 2017 (25.66%) | 673 (25.95%) | 732 (25.20%) | 342 (23.88%) | 270 (28.97%) | |

Abbreviations: SDoH = social determinants of health.
[a] Comparisons across all 4 subphenotypes were performed using $\chi2$ test.

### eTable 16. Logistic regression analyses and Cox regression analyses showing associations between SDoH strata and 60-day mortality risk by subphenotypes in the development cohort

| Subphenotypes | Log odds (SD) | | | Hazard ratio (95% CI) | | |
|---|---|---|---|---|---|---|
| | SDoH stratum H[a] | SDoH stratum M | SDoH stratum L | SDoH stratum H[a] | SDoH stratum M | SDoH stratum L |
| Subphenotype I | - | 0.18 (-0.19 - 0.56) | 0.13 (-0.31 - 0.58) | - | 1.04 (0.74 - 1.47) | 0.99 (0.66 - 1.49) |
| Subphenotype II | - | 0.05 (-0.18 - 0.30) | 0.17 (-0.10 - 0.45) | - | 1.09 (0.89 - 1.35) | 1.13 (0.89 - 1.44) |
| Subphenotype III | - | 0.31 (0 - 0.64) | 0.23 (-0.15 - 0.60) | - | 1.26 (0.96 - 1.66) | 1.14 (0.82 - 1.57) |
| Subphenotype IV | - | 0.15 (-0.20 - 0.50) | 0.40 (0.02 - 0.79)* | - | 1.05 (0.82 - 1.35) | 1.20 (0.92 - 1.57) |

Abbreviations: CI = confidence interval; SD = standard deviation; SDoH = social determinants of health
[a] Reference group.
* P-value < 0.05
